# Comparative performance of plasma pTau181/Aβ42, pTau217/Aβ42 ratios, and individual measurements in detecting brain amyloidosis

**DOI:** 10.1101/2024.12.07.24318640

**Authors:** Sylvain Lehmann, Audrey Gabelle, Marie Duchiron, Germain Busto, Mehdi Morchikh, Constance Delaby, Christophe Hirtz, Etienne Mondesert, Jean-Paul Cristol, Genevieve Barnier-Figue, Florence Perrein, Cédric Turpinat, Snejana Jurici, Karim Bennys, the Alzheimer’s Disease Neuroimaging Initiative (ADNI)

## Abstract

**Background:** Early detection of brain amyloidosis (Aβ+) is crucial for diagnosing Alzheimer’disease (AD) and optimizing patient management, especially in light of emerging treatments. While plasma biomarkers are promising, their combined diagnostic value through specific ratios remains underexplored. In this study, we assess the diagnostic accuracy of plasma pTau isoform (pTau181 and pTau217) to Aβ42 ratios in detecting Aβ+ status.

**Methods:** This study included 423 participants from the multicenter prospective ALZAN cohort, recruited for cognitive complaints. Aβ+ status was determined using cerebrospinal fluid (CSF) biomarkers. The confirmatory cohort comprises 1,176 patient samples from the Alzheimer’s Disease Neuroimaging Initiative (ADNI), with Aβ+ status determined by positron emission tomography (PET) imaging. Plasma biomarkers (pTau181, pTau217, Aβ40, Aβ42) were measured using immunoassays and mass spectrometry, with specific ratios calculated. In the ALZAN cohort, the impact of confounding factors such as age, renal function, ApoE4 status, body mass index, and the delay between blood collection and processing was also evaluated to assess their influence on biomarker concentrations and diagnostic performance. The primary outcome was the diagnostic performance of plasma biomarkers and their ratios for detecting Aβ+ status. Secondary outcomes in the ALZAN cohort included the proportion of patients classified as low, intermediate, or high risk for Aβ+ using a two-cutoff approach.

**Findings:** In ALZAN the pTau181/Aβ42 ratio matched the diagnostic performance of pTau217 (AUC of 0.911 (0.882-0.940) vs. 0.909 (0.879-0.939), P=0.85). The pTau217/Aβ42 ratio demonstrated the highest diagnostic accuracy, with an AUC of 0.927 (0.900-0.954). Both ratios effectively mitigated confounding factors, such as variations in renal function, and were also efficient in identifying Aβ+ status in individuals with early cognitive decline. Diagnostic accuracy of ratios vs. individual measurement was confirmed in the ADNI cohort. In ALZAN, using two-cutoff workflows with pTau217/Aβ42 instead of pTau217 alone reduced the intermediate-risk zone from ∼16% to ∼8%, enhancing stratification for clinical decision-making.

**Interpretation:** The pTau217/Aβ42 ratio demonstrated improved diagnostic performance for detecting Aβ+ compared to individual biomarkers, potentially reducing diagnostic uncertainty. These findings suggest that plasma biomarker ratios could be useful; however, further validation in independent and diverse clinical settings is necessary before broader clinical implementation.

**Funding:** Fondation Research Alzheimer (ALZAN projet), AXA Mécénat Santé (INTERVAL Project), Fondation pour la Recherche Médicale (FRM, team Proteinopathies).

**Research in Context:** *Evidence before this study:* Blood biomarkers of Alzheimer’s disease are receiving increasing attention, as they have great potential for non-invasive detection of the presence of cerebral amyloidosis (Aβ+), one of the key elements in the diagnosis of the disease. Among these biomarkers, the phosphorylated isoforms of the tau protein, principally the pTau217 form, show performances that suggest major interest for clinical use. In addition, a number of studies have demonstrated the value of calculating ratios such as Aβ42/40 or pTau181/Aβ42 in CSF and blood, thereby eliminating the variability associated with pre-analytical or physiological confounding factors, such as renal failure. However, studies directly comparing ratios with individual markers remain limited.

*Added value of this study:* In the prospective real-life ALZAN cohort of cognitively impaired participants we show that plasma pTau181/Aβ42 and pTau217/Aβ42 ratios outperform individual biomarkers for the detection of Aβ+ status established from CSF. This result was confirmed for pTau217/Aβ42 in the ADNI cohort in which detection of Aβ+ status is done with PET. The ratios also show greater resistance to confounding factors such as impaired renal function or delays in sample processing. Finally, by using an approach based on the use of two cutoffs enabling effective population Aβ+ risk stratification, the ratios help to reduce the number of inconclusive situations.

*Implications of all the available evidence:* Taking previous studies into account, our results strengthen the case for using tau and amyloid ratios, and in particular the plasma pTau217/Aβ42 ratio, in the detection of Aβ+ status. These ratios not only improve detection accuracy but also help to take confounding factors into account. Ultimately, they could encourage wider use of routine blood tests, making diagnosis earlier, guiding therapeutic decisions while potentially reducing the use of procedures such as lumbar punctures or PET imaging.

## Introduction

Over the past decade, the contribution of imaging and cerebrospinal fluid (CSF) biomarkers has provided essential elements for the diagnosis of Alzheimer’s disease (AD).^1^ These biomarkers significantly enhance diagnostic confidence when combined with clinical and paraclinical information.^2^ CSF biomarkers are part of the international diagnostic criteria for the disease,^3–5^ enabling more accurate and earlier diagnoses. This is particularly critical for optimizing patient management, especially in the light of emerging treatments, such as anti-amyloid immunotherapies.^6,7^

In north America, cerebral amyloidosis is often detected by amyloid positron emission tomography (PET) imaging,^8^ whereas in Europe, cerebrospinal fluid (CSF) biomarkers, are used to confirm the presence of brain amyloidosis^9^, a hallmark of AD pathology. CSF analysis provides further insights into tau pathology, complementing tau PET, which remains the gold standard for assessing regional tau deposition and distribution.^10,11^ Furthermore, longitudinal studies have established that these biomarkers become positive up to one or two decades before disease onset, during presymptomatic phases.^12^ However, both PET imaging and CSF analysis have limitations: availability, equipment, scalability and associated costs, especially for PET, as well as the invasive nature of lumbar puncture for CSF. These constraints hinder their widespread application, even though the targeted treatments necessitate testing a broader population to evaluate the presence of brain amyloidosis prior to initiating treatment.

The idea of detecting amyloidosis through blood analysis has gained traction in recent years and has undergone significant development and validation. However, additional validation work is still necessary before it can be considered fully suitable for routine clinical application. Early research efforts primarily focused on the detection of amyloid peptides (Aβ).^13^ Detecting Aβ42 and Aβ40 in blood is accurate at around 80% for identifying a positive brain amyloidosis status, called Aβ+.^14^ Total tau protein in blood performs poorly, likely due to its peripheral production.^15^ However, phosphorylated forms of tau protein (pTau) perform much better.^16^ Notably, plasma pTau181, demonstrated an accuracy for Aβ+ of around 80%,^17,18^ close to that of the plasma Aβ42/40 ratio. Following the detection with mass spectrometry of additional pTau isoforms;^19–21^ pTau217 emerged as the biomarker with even higher accuracy for Aβ+, reaching values as high as 90-95% in different studies.^22–26^ It might seem counterintuitive for a tau isoform to indicate amyloidosis and the exact pathophysiological mechanisms behind this observation remain incompletely understood. Nonetheless, the performance of plasma pTau217 is sufficiently powerful to confirm individuals as Aβ+ and contribute to the diagnosis of AD.

A highly promising approach was recently described by Brum et al.^27^, which defines two cutoff values. The lower and upper cutoffs are based on 90% sensitivity and specificity, respectively. Below the 90% sensitivity threshold, Brum et al. (2023) reported the probability of Aβ PET+ to be 42% or less, and above the 90% specificity threshold, they reported the probability of Aβ PET+ to be 70% or more. The effectiveness of this approach is measured by the size of the zone between these two cutoffs, referred to as the “intermediate zone”, which should be minimized. This two cutoffs approach is optimal for tailoring care pathways and even for selecting patients who could benefit from innovative treatments. Immunoassays for pTau217 are now being developed by numerous suppliers, offering highly reliable detection for Aβ+. Head-to-head comparative studies of various assays gave largely equivalent results, with mass spectrometry methods offering a slight edge in accuracy over immunoassays.^16^ These performance levels are suitable for routine use, though additional studies are needed to confirm their robustness in real-world settings. In fact, it has already been established that pre-analytical conditions,^28^ particularly for Aβ peptides, as well as physiological factors, like renal insufficiency,^17,26,29^ or even some drug treatment^30^ can influence concentrations, leading to potential false positives or negatives.

Here we comprehensively evaluate blood Aβ peptides and pTau181 or pTau217 as ratios. By conducting this investigation within the multicentric, real-life ALZAN cohort, and confirming findings in the independent Alzheimer’s Disease Neuroimaging Initiative (ADNI) cohort, we identified the pTau217/Aβ42 ratio as a valuable single metric for Aβ+ detection, providing clinically meaningful high and low cutoffs.

## Methods

### Study populations

This study included participants from the ALZAN multicentre prospective cohort (ClinicalTrials.gov Identifier #NCT05427448). The ALZAN cohort is a clinical study designed to evaluate the diagnostic performance of blood biomarkers for AD. Participants are recruited from memory clinics and neurology departments at the Montpellier, Nîmes, and Perpignan hospitals in France. The inclusion criteria require adults aged 50 to 85 years with cognitive complaints consistent with early-stage AD. Exclusion criteria include significant neurological or psychiatric disorders other than AD that could affect cognition, uncontrolled medical conditions that might interfere with study participation, and recent participation in other clinical trials that could confound results. Sex was self-reported by study participants and used as a covariate in the statistical analysis. Recruitment pathways involve identifying participants through routine consultations at participating centers. Potential candidates undergo preliminary assessments, and those meeting the inclusion criteria are invited to participate. The study protocol has been reviewed and approved by the appropriate ethics committees. All participants provide written informed consent before any study-related procedures are conducted. The consent process includes a detailed presentation of the study’s purpose, procedures, potential risks, and benefits, ensuring that participants understand their involvement is voluntary and that they can withdraw at any time without consequence. Each participant had a lumbar puncture with CSF analysis as part of their routine care.

Two sets of data, originating from the Alzheimer’s Disease Neuroimaging Initiative (ADNI) (www.loni.ucla.edu/ADNI), were used after the agreement of the scientific committee. The first dataset (FNIHBC_BLOOD_BIOMARKER_TRAJECTORIES) relates to the study “Clinical Utility of Blood Biomarker Trajectories” run by the Foundation for the National Institutes of Health (FNIH) Biomarkers Consortium (see acknowledgment). We selected Aβ42, Aβ40 and pTau217 data generated by immunoassays using Lumipulse G®^31^®^31^ and mass spectrometry using C2N Diagnostics methods.^32^.^32^ The second dataset (UCBERKELEY_AMY_6MM) provides the PET amyloid status (cutpoints for Florbetapir (FBP): 20 centiloids (CL); for Florbetaben (FBB): 18 CL) corresponding to the 1176 blood samples from the FNIHBC study. Importantly the mean delay between amyloid PET status and blood collection was 15.6±46.7 days.

### CSF analysis in ALZAN

CSF was collected using standardized protocols for collection, centrifugation, and storage.^33^ CSF Aβ1-42 and Aβ1-40 (referred to here as Aβ42 and Aβ40) were measured using commercially available assays (Fujirebio Europe, Ghent, Belgium) on the Lumipulse G1200 analyzer, following the manufacturer’s protocols. The CSF Aβ42/Aβ40 ratio was used as a biomarker for amyloid pathology, as it accounts for inter-individual differences in total amyloid production and enhances the specificity of Aβ42 as a diagnostic marker.^34^ A lower Aβ42/Aβ40 ratio reflects the preferential aggregation and deposition of Aβ42 in amyloid plaques, leading to its decreased presence in CSF. Participants were therefore categorized as amyloid-positive (Aβ+) or amyloid-negative (Aβ−) based on their CSF Aβ42/Aβ40 ratio, using a threshold set at 7%, which corresponds well to the optimal cutoff identified for detecting amyloid PET status ^35^ as well as AD ^31^.

### Blood Biomarker in ALZAN

Blood samples were collected in EDTA-K2 tubes, the same day as the lumbar puncture and sent to the local clinical laboratory for processing (2000 g × 10 min, RT). The delay between collection and processing in the laboratory (referred to as “blood delay”) was recorded. Plasma aliquots were stored at −80°C in low-binding Eppendorf® LoBind microtubes (Eppendorf, ref 022431064, Hamburg, Germany) until testing. eGFR based on creatinine, age, and sex was calculated using the CKD Epidemiology Collaboration (CKD-EPI) equation, revised in 2021 without inclusion of race.^36^

For biomarker measurement, plasma aliquots were thawed at +4°C, gently homogenized, and centrifuged for 5 minutes at 2000g before analysis. One aliquot was used for pTau217 measurement using commercial kits on the Lumipulse G1200 as reported.^22^ Another aliquot was used for pTau181, Aβ42, Aβ40, and ApoE4 measurement using the Elecsys® RUO (research use only) assays on the e402 Cobas analyzer as described.^37^ All samples were analyzed by staff who were blinded to the clinical data, and all samples had measurable biomarker values. Intra- and inter-run precision, evaluated using pools of clinical samples run across consecutive assays and within a single run, are provided in the supplementary materials, along with the lower limit of quantification (Supplementary table 1).

### Statistical Analyses

The sample size was calculated at the design stage, based on the desired precision of the primary diagnostic accuracy estimates. Specifically, we aimed to estimate sensitivity and specificity with a 95% confidence interval of ±5%, assuming a performance of 85%. Using the standard formula for estimating a proportion with specified precision (n = (Z² × p × (1 − p)) / d²) (where n=sample size, Z = z-score for 95% confidence (1.96), p = expected sensitivity or specificity, d = half-width of the confidence interval), we determined that 196 participants were needed in each group (Aβ+ and Aβ−). Given an anticipated Aβ+ prevalence of 55%, the total required sample size was approximately 356 participants. Allowing for a 5– 10% dropout rate, we targeted up to 440 participants. These calculations were performed using the binDesign() function in the MKmisc package in R. Categorical variables were analyzed as counts and percentages (n, %), while continuous variables, which did not follow a normal distribution as assessed by the Shapiro–Wilk test and illustrated using Q-Q plots (see supplementary data), were reported as medians with interquartile ranges (25th–75th percentiles). The Chi-square test was used to assess group differences for categorical variables because the assumptions for its validity were met: no expected cell count was less than 1, and fewer than 20% of expected cell counts were below 5. Since continuous variables did not follow a normal distribution, the Wilcoxon rank-sum test was used for comparisons between two groups. For comparison with adjustment performed with R, continuous variables were analyzed using linear regression models (lm), and binary variables using logistic regression (glm with family = binomial). Potential confounders were identified based on the modified disjunctive cause criterion, which recommends adjusting for variables that are causal predictors of either the exposure (APOE ε4 status), the outcome (Aβ+ status), or both. Guided by subject-matter knowledge and existing literature, age and sex were identified as such variables and were therefore included as covariates in the adjusted analyses ^38^. We assessed key assumptions of linear regression models: normality of residuals using the Shapiro-Wilk test (shapiro.test), homoscedasticity using the Breusch-Pagan test (bptest from the lmtest package), and linearity via correlation between residuals and fitted values (cor.test). If normality was violated, the dependent variable was log-transformed. If heteroscedasticity was detected, robust standard errors were applied using vcovHC from the sandwich package with coeftest. In case of non-linearity, a natural spline transformation was applied using ns from the splines package (we used a natural cubic spline with 3 degrees of freedom). The performance of individual and ratio biomarkers for Aβ+ detection was assessed using a non-parametric bootstrap resampling to estimate the area under the ROC curve (AUC) and its 95% confidence interval (obtained as the 2.5th and 97.5th percentiles). 1,000 bootstrap iterations were performed using the boot package in R. At each iteration, a new dataset was created by randomly sampling observations with replacement from the original dataset, maintaining the same sample size. Because of this resampling method, each bootstrap sample includes on average about 63.2% of unique observations, while the remaining ∼36.8% may be duplicated or not included at all. AUC was computed for each sample using the pROC package. The distribution of the 1,000 AUC values was summarized using the percentile method: the 2.5th and 97.5th percentiles were reported as the bounds of the 95% confidence interval. Subsequently, pairwise comparisons between AUCs were conducted using DeLong method ^39^ and the resulting p-values were adjusted for multiple comparisons using Benjamini-Hochberg (FDR) procedures. Forest plots of associations between biomarkers and demographic/comorbidity features uses linear regression between the standardized biomarker and each clinical predictor (normalization process applies z-score standardization by subtracting the mean and dividing by the standard deviation for each variable). Because both the outcome and predictors are standardized, the slope from these regressions is numerically equivalent to the Pearson correlation coefficient. The regression beta coefficient and their 95% confidence intervals are displayed in the plots. Only samples with complete data were included in the analysis, ensuring that no biomarker data were missing. All analyses were performed using MedCalc (20·118) and R (R Core Team (2019)) software.

### Ethical approval

All participants gave written informed consent to be part of the study. The ALZAN study was approved by the “Comité de Protection des Personnes Nord Ouest IV” under # 2022-A00565-38. Clinical trial registration: NCT05427448.

## Results

### Participants and biomarker profile

The participants are part of the ALZAN cohort (NCT05427448), a study designed to prospectively and consecutively evaluate the diagnostic performance of blood biomarkers for AD. Of the 433 ALZAN participants initially enrolled, 423 were included in the final analysis, based on the availability of sufficient CSF and blood samples to measure all biomarkers, with no missing biomarker values. Participants were recruited at three memory clinics, consulting for cognitive complains (Supplementary Table 2). The median age at baseline was 71.1 (interquartile range (IQR), 25^th^-75^th^ percentile: 66.8-77.9). The sex ratio was balanced with 197 men to 226 women. 42.8% were APOE ε4 carriers, as determined by the Elecsys immunoassay.^37^ The median MMSE score at recruitment was 23 (IQR 21-26) with the following distribution, subjective cognitive impairment (15.9%), mild cognitive impairment (33.2%), mild dementia (38.1%) and dementia (12.8%). In terms of kidney function, the median estimated glomerular filtration rate (eGFR) was 84.5 (IQR 76.1-94.9) ml/min/1.73m². Based on the CSF Aβ42/40 ratio,^31^, 57.5% of the population was classified as amyloid-positive (Aβ+). Regarding blood sample processing, over 50% of the patients had their blood samples processed the next day or later (Supplementary Figure 1). All plasma measurements for Aβ40, Aβ42, pTau181, and pTau217 were above their respective lower limits of quantification (Supplementary Table 1).

The ADNI cohort includes patients with varying cognitive statuses, ranging from cognitively normal individuals to those with mild cognitive impairment and AD. Participants undergo comprehensive assessments, including neuroimaging (MRI and PET scans), fluid biomarker analysis, and cognitive testing, aimed at advancing AD research. ADNI samples, measured using different assays (see Methods), were selected by the Foundation for the National Institutes of Health (FNIH) Biomarkers Consortium to evaluate the performance of various blood biomarkers. These samples were obtained from individuals who had plasma collected within six months of undergoing an amyloid PET (florbetapir) scan. The median age was 76.4 (70.2-81.4) and the sex ratio was balanced at 50% (Supplementary Table 3).

### Comparison of cohort participant characteristics between Aβ− and Aβ+ groups

Participants were separated into Aβ− and Aβ+ groups based on CSF Aβ42/40 ratio, using a cutoff of 0.07 in ALZAN or on PET amyloid in ADNI (see Methods). Table 1 reports for ALZAN the demographic, clinical, and biomarker readings of the two groups. Key differences include: Aβ+ participants were significantly older than Aβ-participants (median (IQR) 74.9 (70.8-79.1) vs. 69.8 (60.0-76.0); P<0.001), and were less likely to be male (42.0% vs. 52.8%, P = 0.0354). The Aβ+ group had more APOE ε4 carriers (58.4% vs. 21.7%, P<0.001), and Aβ+ participants had lower MMSE scores (median (IQR) 23 (20-25) vs. 25 (22-28); P<0.001). For blood biomarkers, in the Aβ+ group, plasma levels of Aβ42 were significantly lower (median (IQR) 31.9 (28.4-35.8) vs. 38.1 (33.8-42.8);P<0.001. Levels of pTau181 and pTau217 as well as ratio Aβ42 to were significantly higher in the Aβ+ group (Table 1). The fold difference for pTau217 was greater than for pTau181, at 3.63 and 1.84 respectively.

### Using individual plasma biomarkers and their ratios to detect cerebral amyloidosis

We plotted ROC curves for various individual biomarkers and their ratios to evaluate their ability to detect Aβ+ status. Figure1, panels a and b, show in ALZAN the combinations of blood amyloid peptides, with pTau181 and pTau217, respectively, which are then summarised in panel c and Supplementary table 4. In terms of amyloid, Aβ42 performance was intermediate with AUC of 0.739 (0.691-0.787), but ratio, Aβ42/40 performed significantly better with an AUC of 0.84 (0.801-0.879) (P<0.001, note also that the 95% CI do not overlap). For pTau181 (Figure 1a), the biomarker performance was equivalent to that of the Aβ42/40 ratio, with an AUC of 0.857 (0.82–0.894) (P = 0.51). The pTau181/(Aβ42/Aβ40) ratio (AUC of 0.900 (0.869–0.930)) as well as the pTau181/Aβ42 ratio (AUC of 0.911 (0.882–0.940)) were both significantly higher than the Aβ42/40 ratio (P = 0.0039 and P < 0.001, respectively) and pTau181 alone (P < 0.001 for both) (Supplementary Table 4).

**Figure 1.**
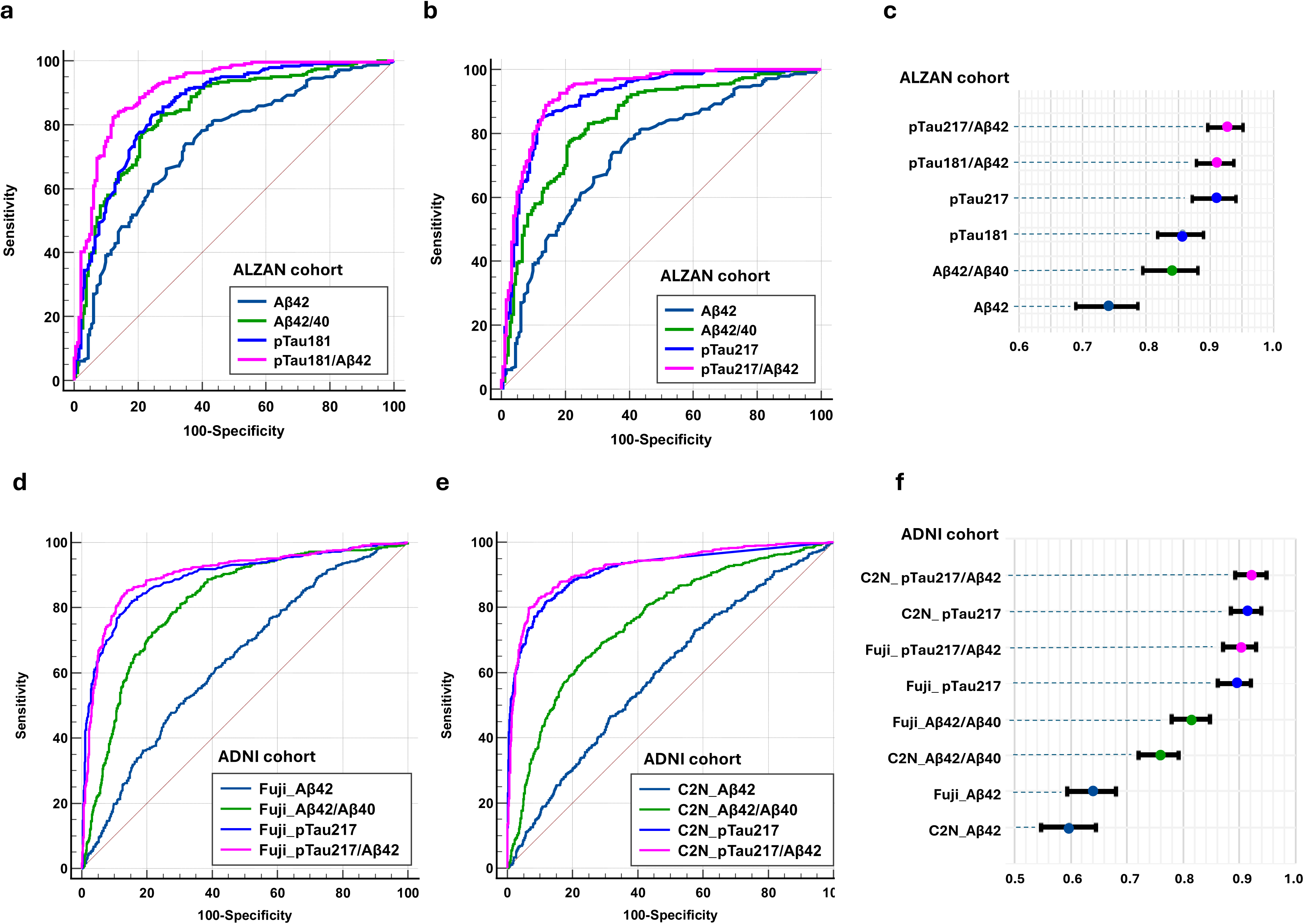
ROC curves of plasma biomarkers according to amyloid status in ALZAN and ADNI. Receiver operating characteristic curve curves for Aβ+ detection in ALZAN (a-c) and ADNI (d-f) using Aβ40, Aβ42, and Aβ42/40 in combination with pTau181 (a) or pTau217 (b,d,e). The corresponding areas under curve (AUC) with 95% confidence intervals are represented in (c) and (f). In ADNI, biomarkers were measured using either an immunoassay approach (Fuji) (d) or with mass spectrometry (C2N) (e). AUC comparisons are reported in Supplementary Table 4 and 5.

The tendency of studies over the last five years has been to assign pTau217 superiority to p181 in detecting amyloid positivity.^25,26,40,41^ Consistently, the pTau217 AUC of 0.909 (0.879-0.939) was higher than for pTau181 (0.857 (0.82-0.894); P<0.001) but equivalent to that of pTau181/Aβ42. As with pTau181 the best pTau217 blood biomarker came from just considering the p217/Aβ42 ratio, which gave the best AUC of all in our hands of 0.927 (0.898-0.950).

Figure 1 also illustrates, in the ADNI cohort, the combinations of blood amyloid peptides with pTau217 measured using either immunoassay (panel d) or mass spectrometry (panel e), which are subsequently summarized in panel f and Supplementary Table 5. In both cases, the Aβ42/40 ratio performed better than Aβ42 alone. pTau217 achieved an AUC of 0.894 (0.874-0.913) for immunoassay detection and 0.914 (0.897-0.931) for mass spectrometry, outperforming the Aβ42/40 ratio. When pTau217 was combined with Aβ42, the AUCs improved further, with mass spectrometry showing the best performance.

We also studied in ALZAN the performance of the pTau217 isoforms, with and without Aβ42, to detect Aβ+, when the population was stratified based on MMSE score (supplementary Figure 2). The pTau217/Aβ42 performed better than pTau217 alone within the early AD subgroup (MMSE >25) (AUC 0.931 (0.867-0.970) vs 0.953 (0.896 to 0.984): P=0.010.

### Association of plasma pTau217 and pTau181 levels with different biomarkers and cohort characteristics

We used in ALZAN linear regression to assess the association of plasma biomarkers with age, body mass index (BMI), blood processing delay, and eGFR (see Methods) (Figure 2, supplementary table 6). We profiled the Aβ-population to focus on the confounding factors influencing biomarker concentration. The association with age was notably significant for Aβ40 (β=1.9, P<0.001) and Aβ42 (β=0.24, P<0.001), but not for Aβ42/40 (β=0.0, P=0.91). Among pTau biomarkers, use alone or in ratios, only pTau217/ Aβ42 was not influenced by age (β=0.0054, P=0.080). Blood delay influenced both Aβ40 (β=−1.42, P<0.001) and Aβ42 (β=−1.43, P<0.001), but not Aβ42/40 (β=0.0002, P=0.29). None of the pTau biomarkers were influenced by blood delay. BMI had no notable influence on biomarkers. Renal function, as assessed by eGFR, was highly impactful, significantly affecting Aβ40 (β=−1.7, P<0.001), Aβ42 (β=−0.25, P<0.001), but not Aβ42/40 (β=−0.0002, P=0.25). Among pTau biomarkers, use alone or in ratios, only pTau217/ Aβ42 was not influenced by eGFR (β=−0.0041, P=0.064).

**Figure 2.**
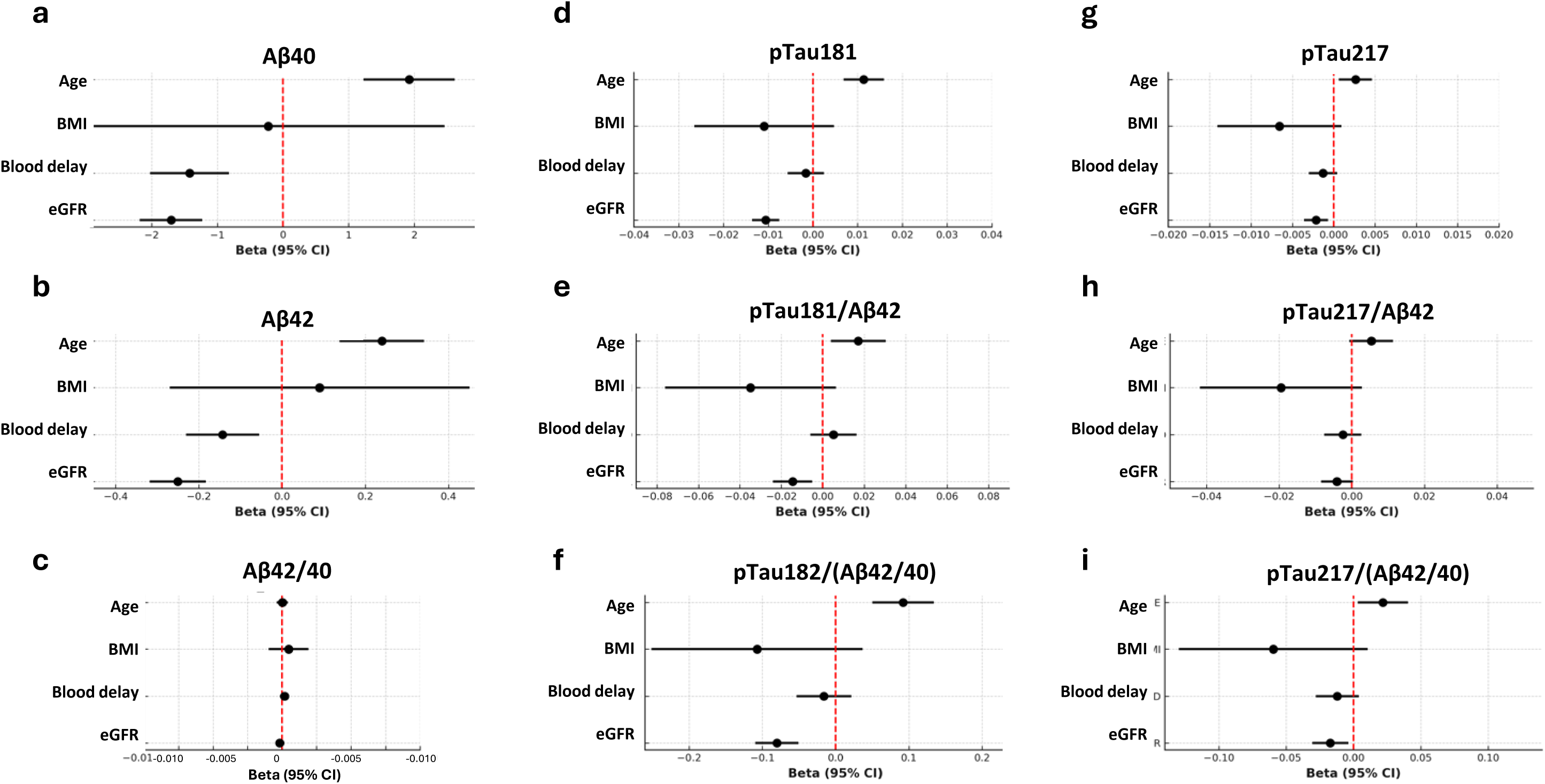
Association of plasma biomarkers and their ratios with ALZAN cohort characteristics. Forest plots of associations in the ALZAN cohort with age, BMI, blood delay and eGFR, using linear regression, for: (a) Aβ40, (b) Aβ42, (c) Aβ42/40, (d) pTau181, (e) pTau181/(Aβ42/40), (f) pTau181/Aβ42, (g) pTau217, (h) pTau217/(Aβ42/40) and (i) pTau217/Aβ42. Regression beta coefficients with 95% confidence intervals are illustrated. Abbreviations: BMI, body mass index; eGFR, estimated glomerular filtration rate.

### Stratifying the population for the risk for Aβ+

Our next objective in ALZAN was to stratify our population for the risk of Aβ+ using pTau181, pTau217, or their ratios with Aβ42, applying a two-cutoff approach.^27^ Instead of selecting the best cutoffs using the commonly used Youden index, we selected cutoffs corresponding to 90% specificity or sensitivity for Aβ+, as recommended by the Global CEO Initiative on AD^42^, in order to remain independent of predictive values, which depend on the prevalence within the population. Nevertheless, we also selected cutoffs to achieve 90% positive or negative predictive values, which are particularly useful in our cohort (see Discussion). The cutoffs, along with their corresponding sensitivity, specificity, and predictive values, are presented in Table 2. The stratification of the population between, low, intermediate and high-risk groups resulting from these two approaches is also reported in Table 2 and illustrated in Figure 3ab. pTau217/Aβ42 shows the smallest intermediate zone, at 7.8% and 8.3% using the sensitivity/specificity and predictive value approaches, respectively. Importantly, these values are reduced by half compared to those of pTau217 alone. Similarly, the pTau181/Aβ42 ratio also improves stratification by reducing the intermediate zone compared to pTau181 alone. The relationship between the selected cutoffs and the ROC curves for pTau217 and pTau217/Aβ42 is shown in Figure 3cd. This figure visually demonstrates how small differences in overall AUC can lead to significant differences when stratifying the population.

**Figure 3.**
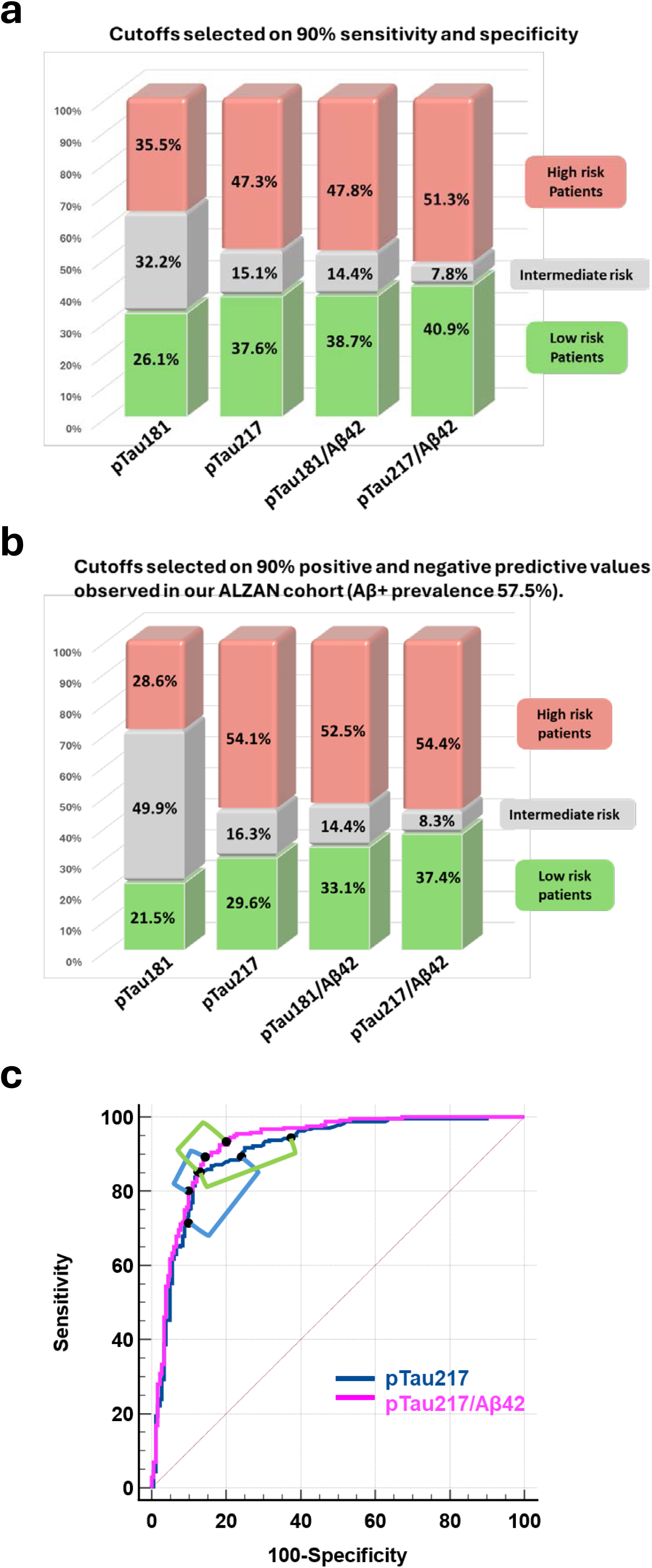
Distribution of the ALZAN population between high-, intermediate-, and low-risk brain Aβ+ groups. Histograms of the ALZAN population, distributed among high-, intermediate-, and low-risk brain amyloidosis groups, are shown as defined by the cut points of pTau181, pTau217, pTau181/Aβ42, and pTau217/Aβ42 (see Table 2). Cutoffs were selected to achieve over 90% specificity/sensitivity (panel a) or over 90% predictive value (with Aβ+ prevalence of 57.5%) (panel b). Panel c: ROC curves for Aβ+ detection using pTau217 (blue) and pTau217/Aβ42 (cyan) were plotted. Respective brackets link the two cut-points used to define low and high predictive risk for Aβ+ using 90% specificity/sensitivity (blue brackets) or 90% predictive value (with Aβ+ prevalence of 57.5%) (green brackets). Note that the brackets are smaller on the pTau217/Aβ42 curve. Abbreviations: ROC: receiver operating characteristic curve.

## Discussion

The data presented here come mainly from the multicentric prospective ALZAN cohort of patients consulting at memory clinics. The rationale of our study was the importance of identifying Aβ+ status, not only to aid in clinical diagnosis but also as a potential triage method for the administration of anti-amyloid therapies. Our study demonstrates that plasma biomarker ratios, particularly plasma pTau217/Aβ42, can identify Aβ+ with high accuracy. Using high and low cutoffs for this single ratio parameter in a two-cutoffs workflow approach reduces the zone of uncertainty by more than half compared to individual phosphorylated tau isoforms. This finding may have important implications for optimizing clinical workflow, particularly in supporting the selection of patients for anti-amyloid treatment, given the observed effect size and direction of association, especially considering that blood biomarkers may be even more accurate than CSF biomarkers.^43^

We compared in ALZAN the ability of plasma biomarkers to detect brain amyloidosis (Aβ+), as defined by the CSF Aβ42/40 ratio. CSF-based amyloid status determination is considered the gold standard in Europe due to the limited use of PET imaging. The concordance between amyloid PET imaging and CSF analyses has been previously established^9^ but is not absolute and may account for some differences in the results obtained between our two cohorts, AZLAN, and ADNI were the amyloid status was defined using PET. We observed performance (AUC values) for Aβ+ detection using pTau181, pTau217, and Aβ42 that are comparable to those reported in the literature.^14,16^ The use of ratios is not novel, as exemplified by the CSF and blood Aβ42/40 ratio, which consistently demonstrates superior performance over Aβ42 alone.^34^ CSF Aβ42/40 reduces false negative/positive since it has the ability to account for Aβ42 levels variation not linked to the amyloid pathology but rather to preanalytical variations (tube type, preservation conditions) or physiological differences.^44–46^ In addition, the fact that CSF Aβ40 levels is increased in AD^10,47^ further enhances the ratio’s diagnostic performance. In blood, the Aβ42/40 ratio additionally mitigates confounding factor such as renal dysfunction, an observation that we initially made^29^ and that we corroborate in our present study. In ALZAN we tracked the interval between blood collection and sample processing, a rarely monitored pre-analytical variable. We could confirm that plasma Aβ levels decreased with the increase of the delay, an observation that was previously made experimentally,^33,48^, and that the use of the Aβ42/40 ratio minimized this bias. Another interesting studied ratio is the pTau181/Aβ42 ratio, which has shown excellent performance in detecting AD in CSF, surpassing both the Aβ42/40 ratio and the individual use of pTau181.^49,50^ Finally, the pTau181/Aβ42 ratio is considered in CSF as the optimal predictor of AD when compared to individual biomarkers.^49,51,52^ The rationale lies, first, in the distinct biological information each biomarker reflects (i.e., amyloid pathology and neurodegeneration^10^), and second, in the mitigation of confounding factors that may affect both biomarkers.

In ALZAN, the plasma pTau181/Aβ42 and pTau217/Aβ42 ratios were associated with a meaningful increase in AUCs for detecting Aβ positivity, compared to pTau measurements alone. The interest of computing the pTau181/Aβ42 ratio in blood was already suggested in recent studies.^53,54^ Substituting Aβ42 with the Aβ42/40 ratio did not yield additional benefit. The improvement of AUCs was particularly notable for the pTau181/Aβ42 ratio and less so for pTau217/Aβ42, likely because pTau217 already exhibits excellent standalone performance. The relatively large size and real-world nature of the ALZAN cohort, with its pre-analytical and clinical variability compared to controlled studies, likely facilitated the demonstration of performance gains for highly effective biomarkers like pTau217. Importantly the superiority of the pTau217/Aβ42 ratio, measured either by immunoassay or mass spectrometry, with regard to Aβ+ detection using PET, was confirmed in the independent ADNI cohort. In ALZAN, the ratio also helped reduce confounding effects related to blood delay and renal function, which however minimally impacted pTau217. This is also reminiscent of the better performance of mass spectrometry-based methods, which also rely on ratios.^32^.^32^ The performance improvement going from pTau217 to pTau217/Aβ42 remained in individuals with MMSE >25 which enhances the potential clinical of the ration for early detection. This fits well with findings that Aβ42 performs better in earlier than in advanced stages of AD.^55,56^.^55,56^

While the AUC increase between pTau217 and pTau217/Aβ42 was statistically significant in both cohorts, the absolute difference was modest. This raises the question of whether the need to measure two biomarkers, each subject to variability and bias,^48^ is justified in routine practice. To complete our evaluation of the potential utility of plasma ratios, we analyzed biomarker performance using two distinct cutoffs for Aβ+ risk stratification.^27^.^27^ We followed the recommendation of the Global CEO Initiative on AD,^42^ by selecting cutoffs with 90% of sensitivity and specificity. Alternatively, we selected cutoffs to achieve 90% positive or negative predictive values in our memory clinic population, as we aim to guide decisions regarding lumbar punctures or imaging while avoiding redundant tests. Using these two approaches, we demonstrate that the pTau217/Aβ42 ratio achieves the best stratification, outperforming pTau217 alone by reducing the proportion of patients in the intermediate (uncertain) category by 50% (from approximately 16% to 8%).

Our study has some limitations. One key limitation is the lack of available neuropathological data. Additionally, comparing cutoffs for one biomarker against others, each with its own predefined thresholds, presents inherent challenges. Additionally, the ALZAN population consists of patients from memory clinics who underwent lumbar puncture for CSF biomarker analysis. Therefore, extrapolating our findings to other settings, particularly primary care where patients may be in the earlier stages of AD, should be approached with caution, even though our results were confirmed in the independent ADNI cohort and remained valid in the ALZAN population with MMSE scores above 25. Of note, two recent studies, one in a community-based, cognitively unimpaired population^57^ and another in non-demented elderly,^58^ have also identified the pTau217/Aβ42 ratio as the best predictor of PET amyloid status. Although these populations differ from ours, which focuses on cognitively impaired patients, making our study original, the consistency of their results further supports and reinforces our conclusions. Two other recently published independent studies in cognitively impaired participants.^22,59^ confirmed the excellent performance of pTau217and the pTau217/Aβ42 ratio, but without highlighting the superior performance of the latter or exploring its relationship with confounding factors. This further strengthens the rationale for using ratios.

In conclusion, this study demonstrated that plasma biomarker ratios, specifically pTau217/Aβ42 and pTau181/Aβ42, significantly enhance the accuracy of Aβ+ detection compared to individual biomarkers, including in the early stages of AD. The performance of pTau181/Aβ42 was comparable to that of pTau217, while pTau217/Aβ42 enabled a two-fold reduction in the intermediate zone during a two-cutoff stratification of Aβ+ risk. These findings highlight the clinical utility of such ratios in stratifying patients for diagnosis and treatment decisions within memory clinics, potentially reducing unnecessary procedures and improving early-stage detection. However, further validation in diverse clinical settings is necessary to confirm these results.

## Data Availability

Data and informed consent forms are available upon request (CHU Montpellier). Requests will be considered by each study investigator, based on the information provided by the requester, regarding the study and analysis plan. If the use is appropriate, a data sharing agreement will be put in place before distributing a fully de-identified version of the dataset, including the data dictionary used for analysis with individual participant data.

## Contributors

S.L., C.D., G.B. and K.B. have accessed and verified the underlying data. They take responsibility for the integrity of the data and the accuracy of the data analysis.

Concept and design: S.L, A.G, K.B.

Acquisition of biological data: M.D, M.M.

Analysis, or interpretation of data: All authors.

Drafting of the manuscript: S.L.

Critical revision of the manuscript for important intellectual content: All authors.

Statistical analysis: S.L, G.B.

Obtained funding: S.L.

All authors had full access to the data and contributed to revision and editing of the manuscript. They read and approved the final version of the manuscript

## Acknowledgement

The work was funded by the Fondation Research Alzheimer (ALZAN projet), AXA Mécénat Santé (INTERVAL Project) and the Fondation pour la Recherche Médicale (FRM, team Proteinopathies).

Data collection and sharing for this project was also funded by the Alzheimer’s Disease Neuroimaging Initiative (ADNI) (National Institutes of Health Grant U01 AG024904) and DOD ADNI (Department of Defence award number W81XWH-12-2-0012). ADNI is funded by the National Institute on Aging, the National Institute of Biomedical Imaging and Bioengineering, and through generous contributions from the following: AbbVie, Alzheimer’s Association; Alzheimer’s Drug Discovery Foundation; Araclon Biotech; BioClinica, Inc.; Biogen; Bristol-Myers Squibb Company; CereSpir, Inc.; Cogstate; Eisai Inc.; Elan Pharmaceuticals, Inc.; Eli Lilly and Company; EuroImmun; F. Hoffmann-La Roche Ltd and its affiliated company Genentech, Inc.; Fujirebio; GE Healthcare; IXICO Ltd.; Janssen Alzheimer Immunotherapy Research & Development, LLC.; Johnson & Johnson Pharmaceutical Research & Development LLC.; Lumosity; Lundbeck; Merck & Co., Inc.; Meso Scale Diagnostics, LLC.; NeuroRx Research; Neurotrack Technologies; Novartis Pharmaceuticals Corporation; Pfizer Inc.; Piramal Imaging; Servier; Takeda Pharmaceutical Company; and Transition Therapeutics. The Canadian Institutes of Health Research is providing funds to support ADNI clinical sites in Canada. Private sector contributions are facilitated by the Foundation for the National Institutes of Health (www.fnih.org). The grantee organization is the Northern California Institute for Research and Education, and the study is coordinated by the Alzheimer’s Therapeutic Research Institute at the University of Southern California. ADNI data are disseminated by the Laboratory for Neuro Imaging at the University of Southern California. We would like to thank Dr. Susan M. Landau (University of California, Berkeley, USA), for her help in using UCBERKELEY amyloid PET datasets. Acknowledgement to the Foundation for the National Institutes of Health (FNIH) Biomarkers Consortium “Plasma Aβ as a Predictor of Amyloid Positivity in Alzheimer’s Disease” for using their results through ADNI. This study was made possible through a public-private partnership managed by the FNIH and funded by AbbVie Inc., Alzheimer’s Association®, Diagnostics Accelerator at the Alzheimer’s Drug Discovery Foundation, Biogen MA Inc., Janssen Research & Development, LLC, Takeda Pharmaceutical Company Limited. The Project Investigators have not participated in reviewing the data analysis or content of the manuscript.

