## Supplementary material for "Comparative performance of plasma pTau181/Aβ42, pTau217/Aβ42 ratios, and individual measurements in detecting brain amyloidosis"

### Supplementary table 1: Assay analytical characteristics

| | Intra-assay CV, % | Inter-assay CV, % | LLOQ, pg/mL |
|---|---|---|---|
| *pTau217 (Lumipulse)* | 3.4 | 9 .4 | 0.03 |
| *pTau181 (Elecsys)* | 2.5 | 3.7 | 0.30 |
| *Aβ40 (Elecsys)* | 0.9 | 7.1 | 10.0 |
| *Aβ42 (Elecsys)* | 3.2 | 4.5 | 0.668 |

Abbreviation: CV: coefficient of variation, LLOQ : lower limit of quantification.

### Supplementary table 2: ALZAN population

| ***Variable*** | ***ALZAN** (n=423)* | ***ALZAN** (n=423)* | **Aβ-** (n=180) | **Aβ+** (n=243) |
|---|---|---|---|---|
| *Age (years)* | 73.6 (66.8 - 77.9) | 71.1 (10.4) | 66.9 (12.6) | 74.2 (7.0) |
| *Men (n, %)* | 197, 46.6% | 197, 46.6% | 95, 52.8% | 102, 42.0% |
| *BMI (kg/m2)* | 24.72 (21.77 - 26.99) | 24.72 (4.19) | 25.64 (4.29) | 24.13 (4.03) |
| *1 or 2 APOE4 alleles (n, %)* | 181, 42.8% | 181, 42.8% | 39, 21.7% | 142, 58.4% |
| *MMSE* | 23 (21 - 26) | 22.9 (4.6) | 24.6 (4.0) | 22.0 (4.6) |
| *eGFR (mL/min/1.73m2)* | 88.1 (76.1 - 94.9) | 84.6 (16.1) | 86.0 (17.5) | 83.5 (14.9) |
| *Blood delay (hours)* | 3.8 (1.50 - 20.00) | 10.11 (14.84) | 11.76 (15.19) | 8.87 (14.49) |
| *Aβ40 (pg/mL)* | 293 (268 - 325) | 298.2 (54.3) | 294.0 (59.2) | 301.4 (50.2) |
| *Aβ42 (pg/mL)* | 34.2 (30.0 - 39.3) | 35.0 (8.5) | 38.7 (8.7) | 32.3 (7.2) |
| *Aβ42/40* | 0.1140 (0.1030 - 0.1310) | 0.119 (0.0275) | 0.134 (0.031) | 0.1070 (0.0175) |
| *pTau181 (pg/mL)* | 1.150(0.759 - 1.588) | 1.283 (0.736) | 0.866 (0.450) | 1.592 (0.755) |
| *pTau217 (pg/mL)* | 0.263 (0.106 - 0.569) | 0.406 (0.415) | 0.162 (0.237) | 0.588 (0.425) |
| *pTau181/Aβ42 ** | 3.349 (2.092 - 5.033) | 3.936 (2.646) | 2.289 (1.212) | 5.156 (2.758) |
| *pTau217/Aβ42 ** | 0.788 (0.282 - 1.898) | 1.285 (1.398) | 0.435 (0.641) | 1.914 (1.472) |

Values are presented for continuous variables as median (25th-75th percentiles) for the first column and as mean (SD) for the other columns. Abbreviations: APOE, apolipoprotein E; BMI, body mass index; MMSE, Mini–Mental State Examination; eGFR, estimated glomerular filtration rate. * The values of the ratio have been multiplied by 100 for better readability.

### Supplementary table 3: ADNI population

| Variable | ADNI (n=1176) | PET Aβ – (n=659) | PET Aβ + (n=516) | P | P$ |
|---|---|---|---|---|---|
| Age (years) | 76.4 (70.2 - 81.4) | 74.1 (68.9 - 79.7) | 77.7 (72.4 - 82.9) | <0.0001 | / |
| Men (n,%) | 588, 50.0% | 324, 49.4% | 264, 51.2% | 0.64 | / |
| Fuji Aβ42 (pg/mL) | 26.23 (22.45 - 29.90) | 26.92 (23.6 - 31.4) | 24.16 (21.31 - 28.11) | <0.0001 | <0.0001 |
| Fuji pTau217 (pg/mL) | 0.2350 (0.0765 - 0.2820) | *0.0880 (0.0565 - 0.125)* | 0.2970 (0.1800 - 0.4600) | <0.0001 | <0.0001 |
| Fuji pTau217//Aβ42 | 0.0121 (0.00281 - 0.0113) | *0.317 (0.2105 - 0.4617)* | 1.2111 (0.7424 - 1.9499) | <0.0001 | <0.0001 |
| C2N Aβ42 (pg/mL) | 39.743 (34.061 - 44.408) | 40.036 (34.9475 - 46.087) | 37.752 (32.809 - 42.273) | <0.0001 | <0.0001 |
| C2N pTau217 (pg/mL) | 2.8370 (0.6500 - 3.5860) | 0.6500 (0.6500 - 1.6140) | 3.913 (2.486 - 6.094) | <0.0001 | <0.0001 |
| C2N pTau217/Aβ42 | 0.0756 (0.0182 - 0.0982) | 1.9675 (1.6039 - 4.0245) | 10.5157 (6.8814 - 15.9653) | <0.0001 | <0.0001 |

Values are presented for (n) samples of the total cohort and for amyloid-negative (Aβ–) and amyloid-positive (Aβ+) groups. As continuous variables did not follow a normal distribution, as assessed using the Shapiro-Wilk test, they were presented as median (25th-75th percentiles) and the Wilcoxon rank-sum test (P) was used to compare negative Aβ– and Aβ+ groups. Additionally, adjusted p-values ($) were obtained using regression models adapted to data's normality, homoscedasticity, and linearity assumptions, with age and sex included as covariates (see Methods). For categorical variables, percentages are provided, and Chi-square tests were conducted to assess group differences.

### Supplementary table 4: AUCs of ROC curves for Aβ+ detection in ALZAN

| Variable | AUC (95% CI) | Aβ42 | Aβ42/40 | pTau181 | pTau217 | pTau181/(Aβ42/40) | pTau217/(Aβ42/40) | pTau181/Aβ42 |
|---|---|---|---|---|---|---|---|---|
| **Aβ42** | 0.739 (0.691 - 0.787) | / | | | | | | |
| **Aβ42/40** | 0.840 (0.801 - 0.879) | < 0.0001 | / | | | | | |
| **pTau181** | 0.857 (0.82 - 0.894) | < 0.0001 | 0.51 | / | | | | |
| **pTau217** | 0.909 (0.879 - 0.939) | < 0.0001 | 0.0027 | < 0.0001 | / | | | |
| **pTau181/(Aβ42/40)** | 0.900 (0.869 - 0.930) | < 0.0001 | 0.0039 | < 0.0001 | 0.44 | / | | |
| **pTau217/(Aβ42/40)** | 0.922 (0.894 - 0.95) | < 0.0001 | < 0.0001 | < 0.0001 | < 0.0001 | 0.040 | / | |
| **pTau181/Aβ42** | 0.911 (0.882 - 0.940) | < 0.0001 | < 0.0001 | < 0.0001 | 0.85 | 0.13 | 0.31 | / |
| **pTau217/Aβ42** | 0.927 (0.900 - 0.954) | < 0.0001 | < 0.0001 | < 0.0001 | < 0.0001 | 0.025 | 0.10 | < 0.0001 |

Column 1 lists the tested biomarkers and their ratios. Column 2 gives AUC values with their 95% CI obtained via bootstrapping. for Aβ+ detection in the ALZAN cohort. The right-hand part show p-values of Delong comparison adjusted for multiple comparison using the Benjamini-Hochberg procedure.

Abbreviations: AUC: area under the curve; ROC: Receiver Operation Curve; CI: confidence interval.

### Supplementary table 5: AUCs of ROC curves for Aβ+ detection in ADNI

| Variable | AUC (95% CI) | C2N_Aβ42 | Fuji_Aβ42 | C2N_Aβ42/40 | Fuji_ Aβ42/40 | Fuji_pTau217 | Fuji_pTau217/Aβ42 | C2N_pTau217 |
|---|---|---|---|---|---|---|---|---|
| ***C2N_Aβ42*** | 0.597 (0.569 - 0.625) | / | | | | | | |
| ***Fuji_Aβ42*** | 0.638 (0.606 - 0.669) | < 0.0001 | / | | | | | |
| ***C2N_Aβ42/40*** | 0.759 (0.731 - 0.787) | < 0.0001 | < 0.0001 | / | | | | |
| ***Fuji_ Aβ42/40*** | 0.816 (0.791 - 0.841) | < 0.0001 | < 0.0001 | < 0.0001 | / | | | |
| ***Fuji_pTau217*** | 0.894 (0.874 - 0.913) | < 0.0001 | < 0.0001 | < 0.0001 | < 0.0001 | / | | |
| ***Fuji_pTau217/Aβ42*** | 0.902 (0.883 - 0.921) | < 0.0001 | < 0.0001 | < 0.0001 | < 0.0001 | 0.030 | / | |
| ***C2N_pTau217*** | 0.914 (0.897 - 0.931) | < 0.0001 | < 0.0001 | < 0.0001 | < 0.0001 | 0.0037 | 0.10 | / |
| ***C2N_pTau217/Aβ42*** | 0.923 (0.907 - 0.939) | < 0.0001 | < 0.0001 | < 0.0001 | < 0.0001 | < 0.0001 | 0.0049 | 0.0059 |

Column 1 lists the tested biomarkers and their ratios. Column 2 gives AUC values with bootstrap with 95% CI for Aβ+ detection in the ADNI cohort. The right-hand part show p-values of Delong comparison adjusted for multiple comparison using the Benjamini-Hochberg procedure.

Abbreviations: AUC: area under the curve; ROC: Receiver Operation Curve; CI: confidence interval.

### Supplementary table 6: Association between biomarkers and confounding factors in ALZAN

| Biomarker | Factor | Regression beta coefficient (95% CI) | P values |
|---|---|---|---|
| Aβ40 | Age (years) | 1.9222 (1.2284 - 2.616) | <0.0001 |
| | Blood delay | -1.423 (-2.0233 - -0.8227) | <0.0001 |
| | BMI (kg/m2) | -0.2274 (-2.9156 - 2.4607) | 0.87 |
| | eGFR | -1.7045 (-2.1818 - -1.2272) | <0.0001 |
| Aβ42 | Age (years) | 0.2399 (0.1383 - 0.3415) | <0.0001 |
| | Blood delay | -0.143 (-0.2312 - -0.0549) | 0.0016 |
| | BMI (kg/m2) | 0.0902 (-0.2701 - 0.4505) | 0.62 |
| | eGFR | -0.2509 (-0.3184 - -0.1833) | <0.0001 |
| Aβ4240 | Age (years) | 0.00 (-0.0004 - 0.0004) | 0.91 |
| | Blood delay | 0.0002 (-0.0002 - 0.0005) | 0.29 |
| | BMI (kg/m2) | 0.0005 (-0.001 - 0.0019) | 0.52 |
| | eGFR | -0.0002 (-0.0005 - 0.0001) | 0.25 |
| pTau181 | Age (years) | 0.0114 (0.0069 - 0.0158) | <0.0001 |
| | Blood delay | -0.0016 (-0.0057 - 0.0025) | 0.43 |
| | BMI (kg/m2) | -0.0109 (-0.0265 - -0.0047) | 0.17 |
| | eGFR | -0.0106 (-0.0137 - -0.0075) | <0.0001 |
| pTau181/(Aβ42/40) | Age (years) | 0.0921 (0.0501 - 0.1341) | <0.0001 |
| | Blood delay | -0.016 (-0.0533 - -0.0213) | 0.40 |
| | BMI (kg/m2) | -0.1073 (-0.2514 - -0.0368) | 0.14 |
| | eGFR | -0.0802 (-0.1098 - -0.0506) | <0.0001 |
| pTau181/Aβ42 | Age (years) | 0.0171 (0.0039 - 0.0304) | 0.011 |
| | Blood delay | 0.0052 (-0.006 - 0.0163) | 0.36 |
| | BMI (kg/m2) | -0.0349 (-0.0762 - 0.0063) | 0.096 |
| | eGFR | -0.0146 (-0.0241 - 0.005) | 0.0029 |

| Biomarker | Factor | Regression beta coefficient (95% CI) | P values |
|---|---|---|---|
| pTau217 | Age (years) | 0.0026 (0.0006 - 0.0046) | 0.011 |
| | Blood delay | -0.0013 (-0.003 - 0.0004) | 0.14 |
| | BMI (kg/m2) | -0.0066 (-0.0141 - 0.0009) | 0.086 |
| | eGFR | -0.0021 (-0.0036 - -0.0007) | 0.0038 |
| pTau217 - (Aβ42 - 40) | Age (years) | 0.0218 (0.0032 - 0.0405) | 0.022 |
| | Blood delay | -0.0121 (-0.028 - 0.0038) | 0.14 |
| | BMI (kg/m2) | -0.0596 (-0.1296 - 0.0105) | 0.095 |
| | eGFR | -0.0172 (-0.0307 - -0.0037) | 0.013 |
| pTau217 - Aβ42 | Age (years) | 0.0054 (-0.0007 - 0.0114) | 0.080 |
| | Blood delay | -0.0024 (-0.0075 - 0.0027) | 0.35 |
| | BMI (kg/m2) | -0.0195 (-0.0418 - 0.0028) | 0.086 |
| | eGFR | -0.0041 (-0.0085 - 0.0002) | 0.064 |

The table provides the regression beta coefficient and the significance level of the association between biomarker values and eGFR. These data are illustrated in Figure 2. Abbreviation: eGFR, estimated glomerular filtration rate; CI: confidence interval.

**Supplementary figure 1**

**Illustration of the delay between blood collection and processing.**

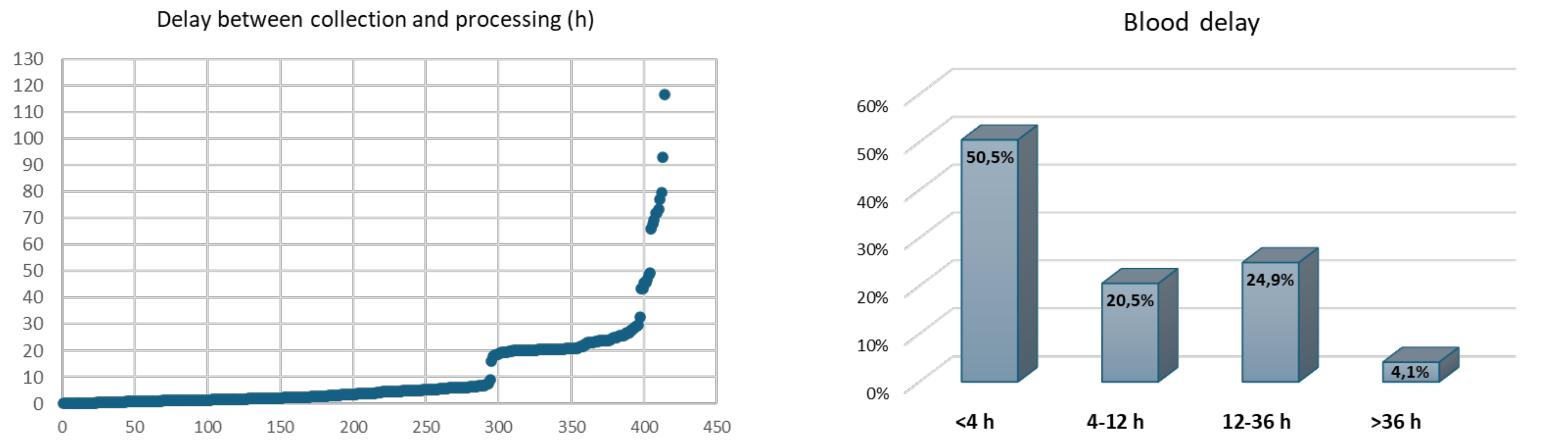


On the left, the delay in hours for the 423 blood samples of the ALZAN cohort is plotted in increasing order. On the right, histograms show the distribution of delay times across four categories: <4h. 4-12h. 12-36h, and >36h. The percentage of the population in each category is also indicated on the bars.

**Supplementary figure 2**

**ROC curves of Aβ42, pTau217, and pTau217/Aβ42 in the ALZAN population stratified by MMSE.**

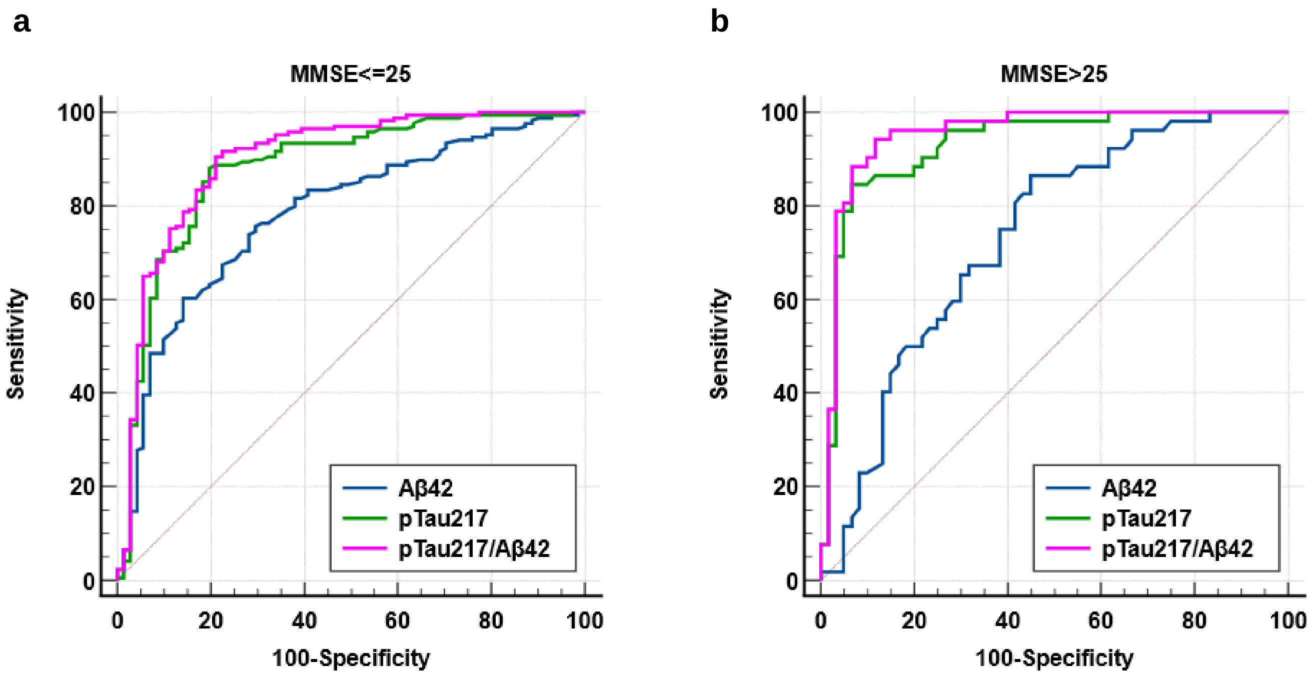


Panel a: AUCs (95% CI) in MMSE<=25 ALZAN population are for Aβ42, pTau217, and pTau217/Aβ42: 0.783 (0.726 to 0.834), 0.879 (0.831 to 0.918) and 0.900 (0.855 to 0.935), respectively. Pairwise p-values comparison using Delong method are: Aβ42 vs. pTau217: 0.0179; Aβ42 vs. pTau217/Aβ42: 0.0019 and pTau217 vs. pTau217/Aβ42: 0.008. Panel b: AUCs (95% CI) in MMSE>25 ALZAN population are for Aβ42, pTau217 and pTau217/Aβ42: 0.732 (0.640 to 0.811), 0.931 (0.867 to 0.970) and 0.953 (0.896 to 0.984), respectively. Pairwise-p values comparison using Delong method are: Aβ42 vs. pTau217: 0.0003; Aβ42 vs. pTau217/Aβ42: <0.0001 and pTau217 vs. pTau217/Aβ42: 0.0101.

Abbreviations: ROC: receiver operating characteristic curve; MMSE. Mini–Mental State Examination.

**Supplementary Q-Q plots**

**Q-Q plot of the ALZAN variables in the Amyloid negative (A(-) and positive A(+) groups**

Legend:
X-axis: Theoretical Quantiles,
Y-axis: Sample Quantiles

Q−Q plot: AGE A (-)

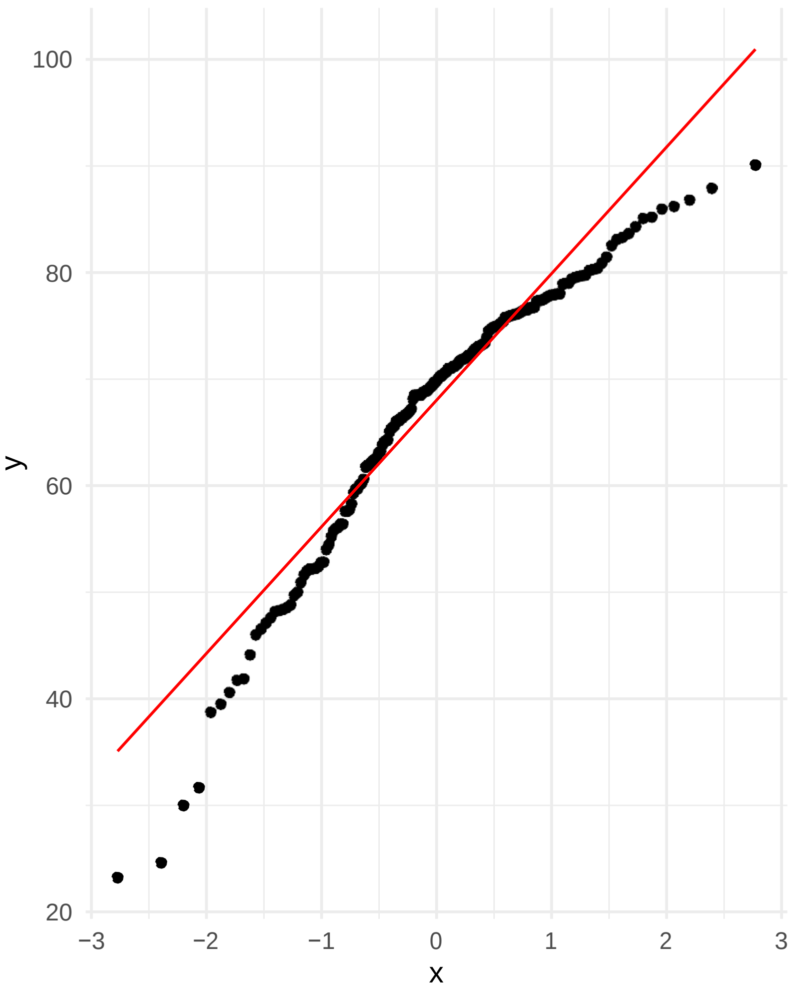


Q−Q plot: AGE A (+)

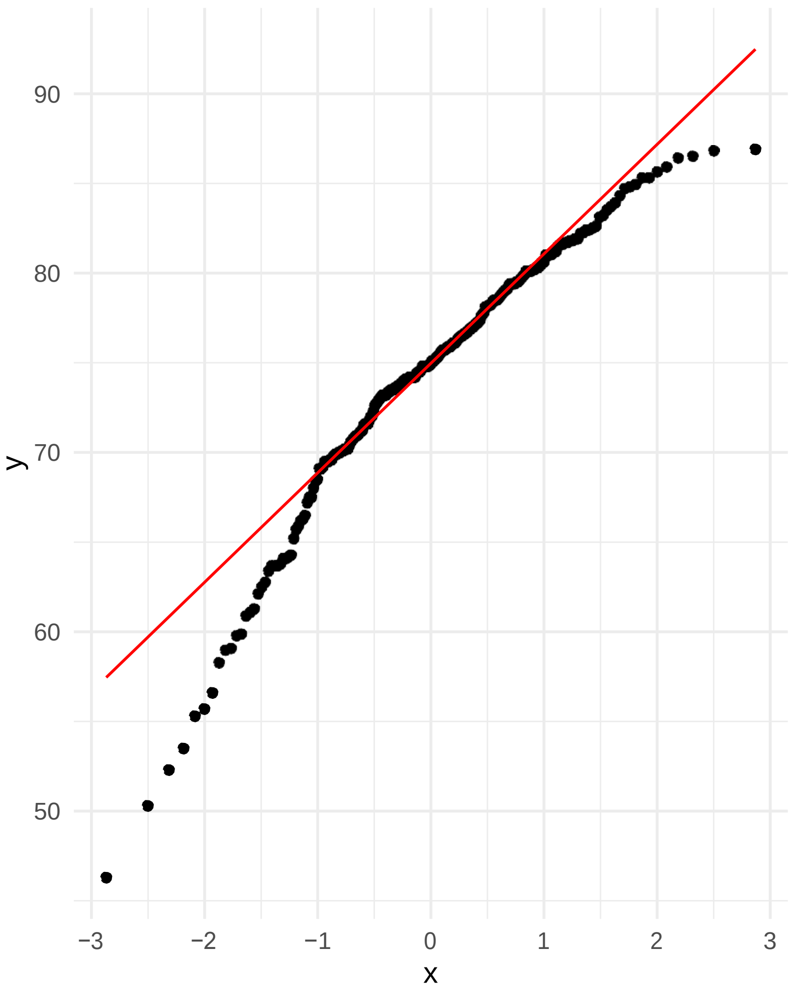


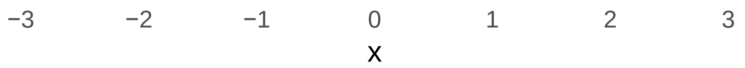


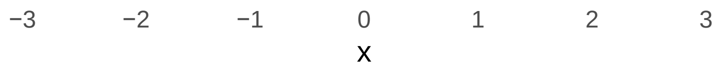

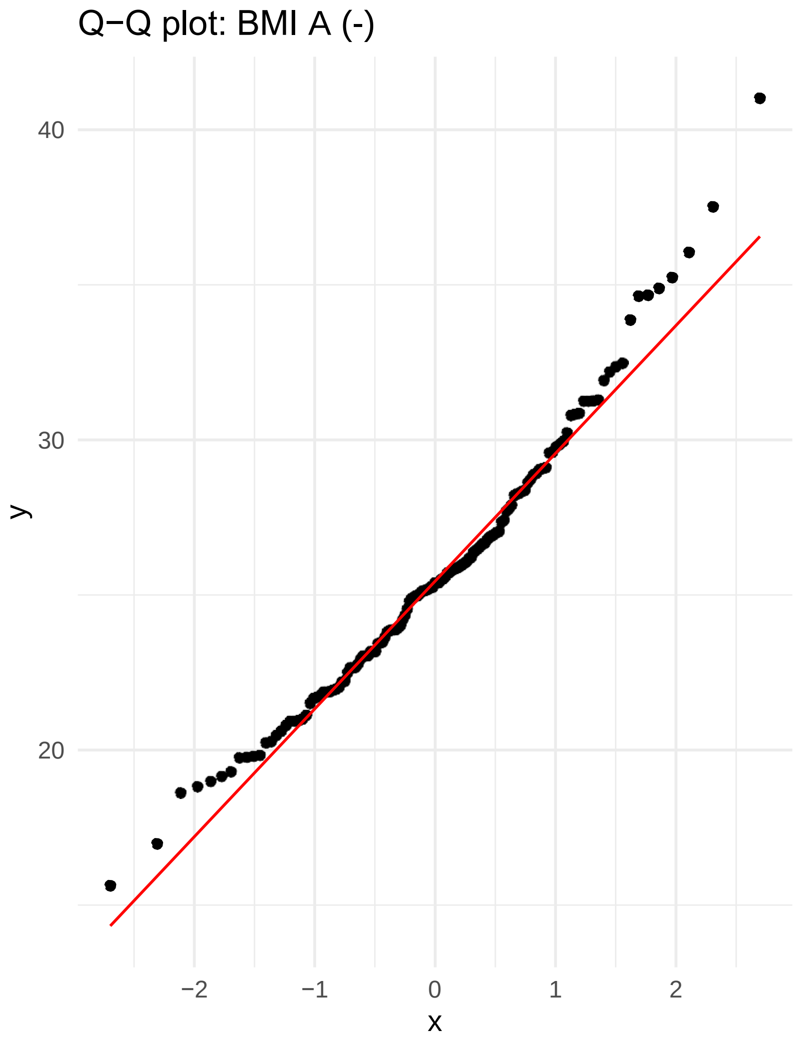
Q−Q plot: BMI A (-)
40
30
20
y
−2
−1
0
1
2
x

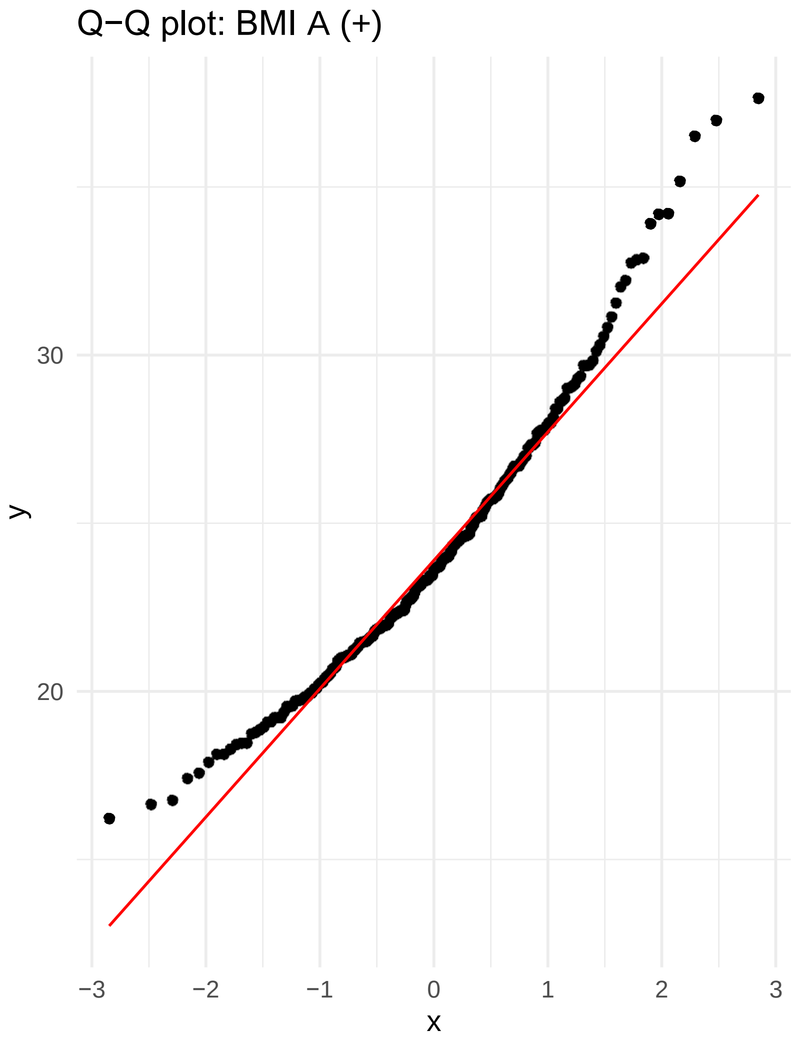
Q−Q plot: BMI A (+)
30
20
y
−3
−2
−1
0
1
2
3
x

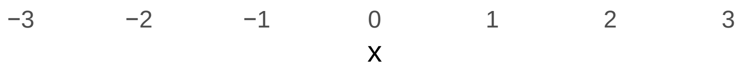
−3
−2
−1
0
1
2
3
x

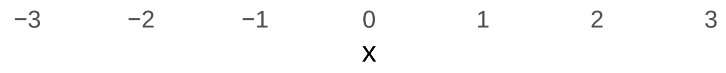
−3
−2
−1
0
1
2
3
x

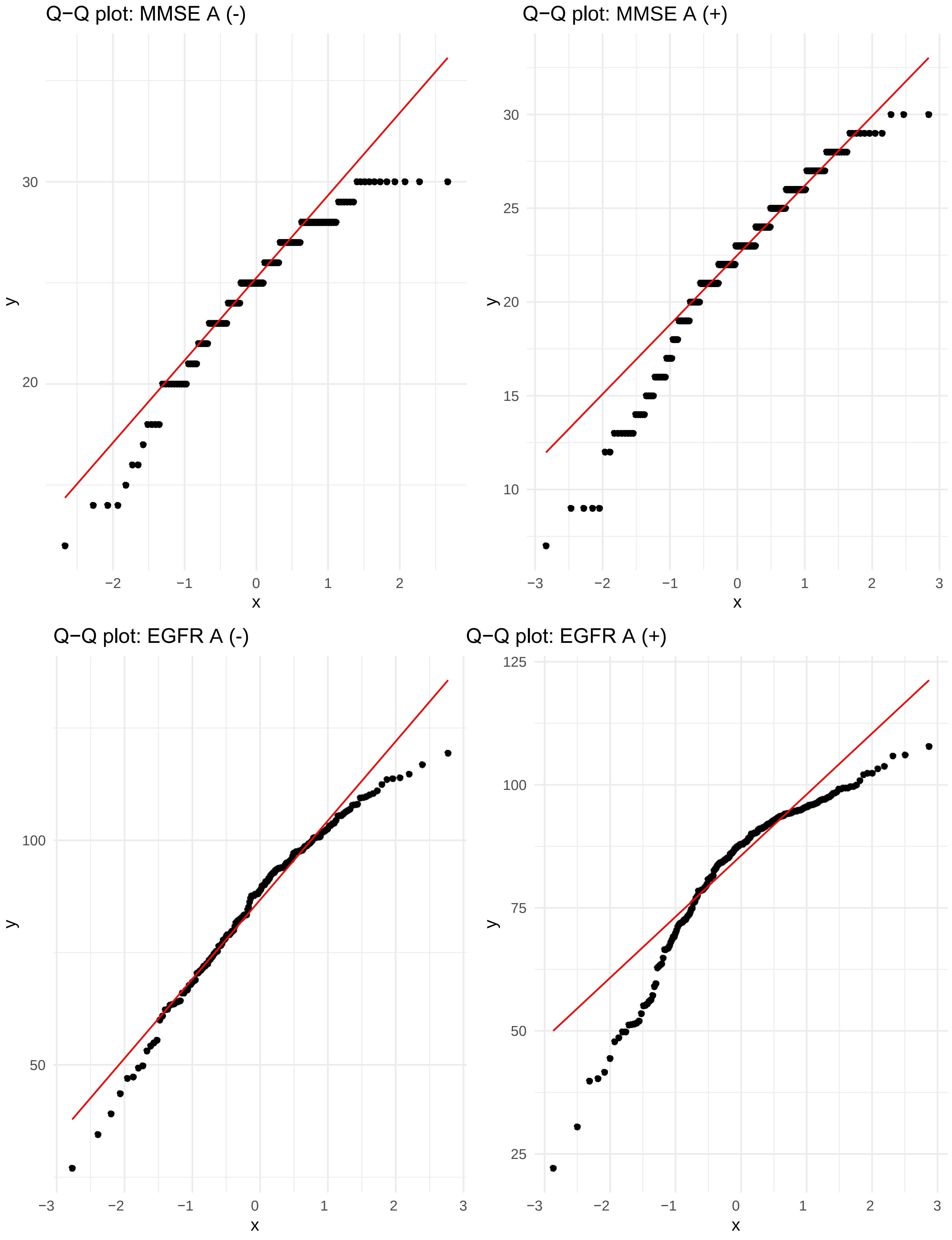
Q−Q plot: MMSE A (-)
30
20
y
−2
−1
0
1
2
x
Q−Q plot: MMSE A (+)
30
25
20
15
10
y
−3
−2
−1
0
1
2
3
x
Q−Q plot: EGFR A (-)
100
50
y
−3
−2
−1
0
1
2
3
x
Q−Q plot: EGFR A (+)
125
100
75
50
25
y
−3
−2
−1
0
1
2
3
x

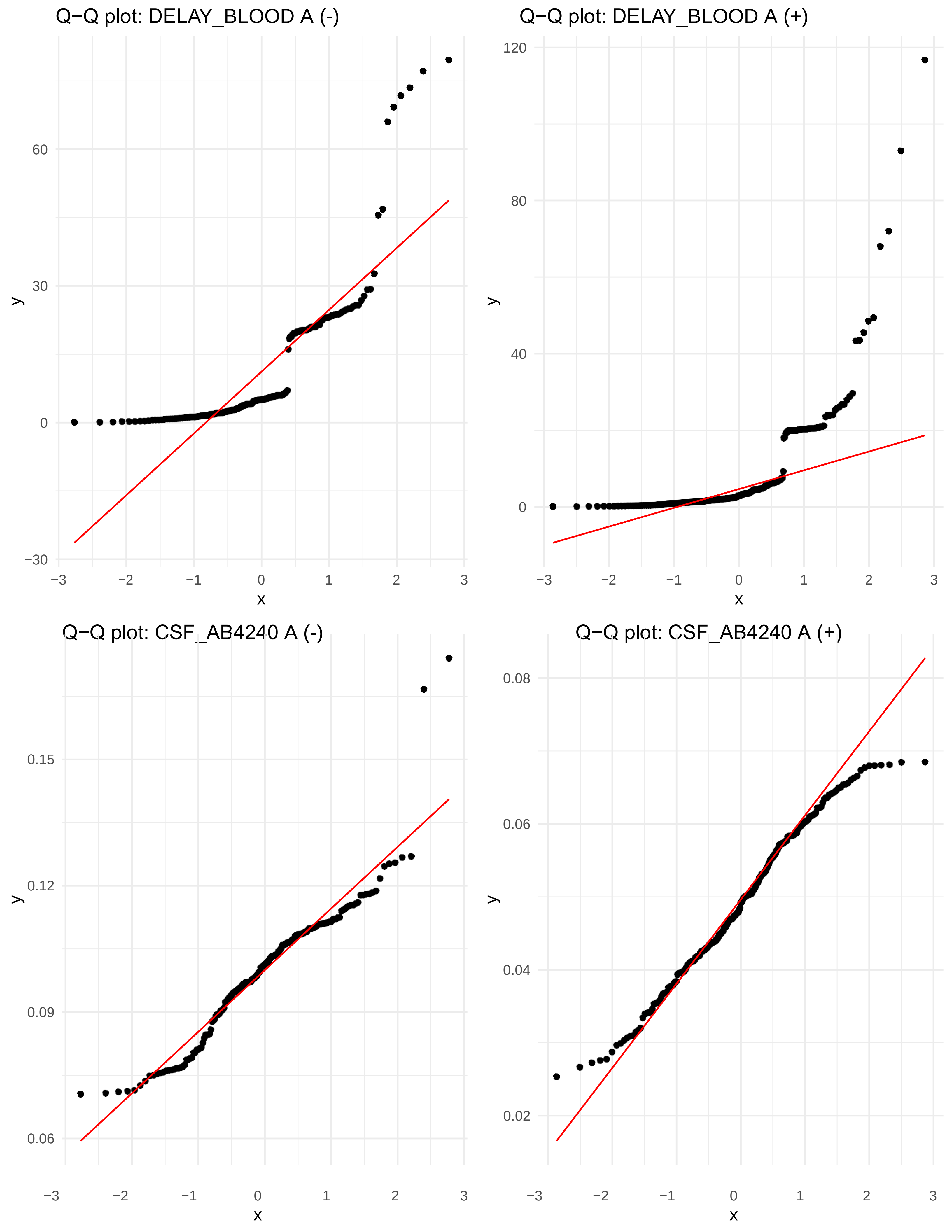

Q−Q plot: DELAY_BLOOD A (-)
Q−Q plot: DELAY_BLOOD A (+)
Q−Q plot: CSF_AB4240 A (-)
Q−Q plot: CSF_AB4240 A (+)
x
y
−30
0
30
60
40
80
120
0.06
0.09
0.12
0.15
0.02
0.04
0.06
0.08
−3
−2
−1
0
1
2
3

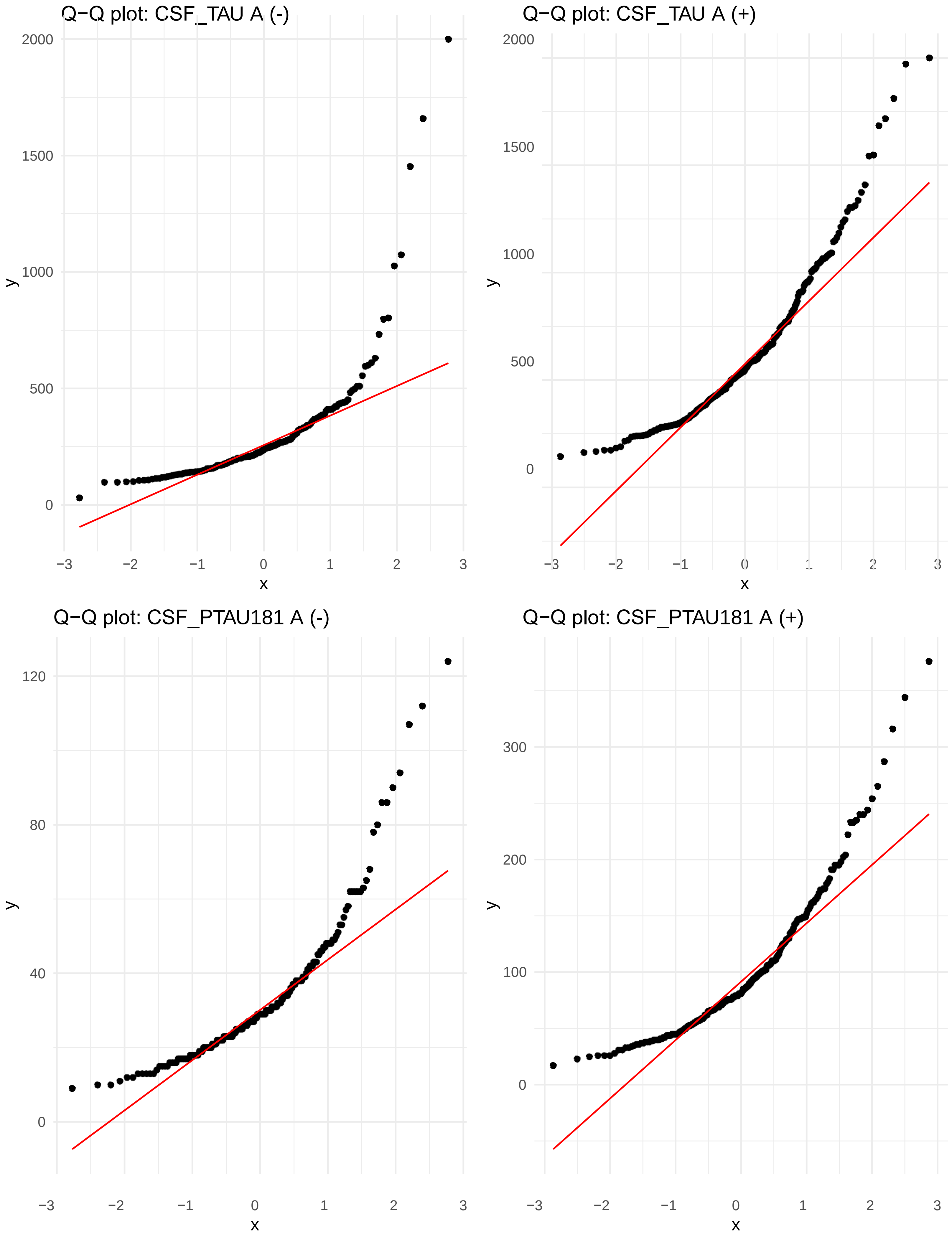

Q−Q plot: CSF_TAU A (-)
Q−Q plot: CSF_TAU A (+)
Q−Q plot: CSF_PTAU181 A (-)
Q−Q plot: CSF_PTAU181 A (+)
y
x

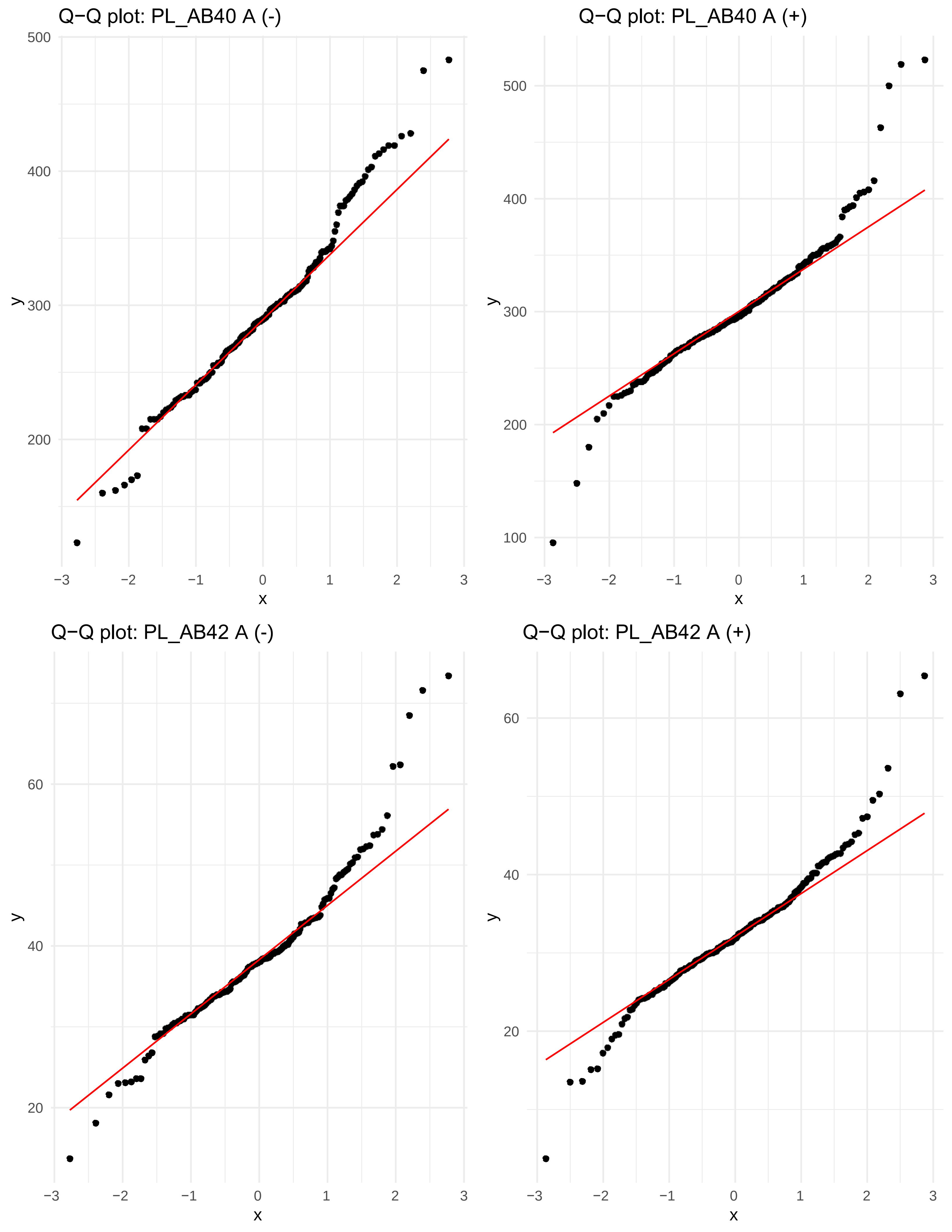
Q−Q plot: PL_AB40 A (-)
Q−Q plot: PL_AB40 A (+)
Q−Q plot: PL_AB42 A (-)
Q−Q plot: PL_AB42 A (+)
y
x
500
400
300
200
100
60
40
20
−3
−2
−1
0
1
2
3

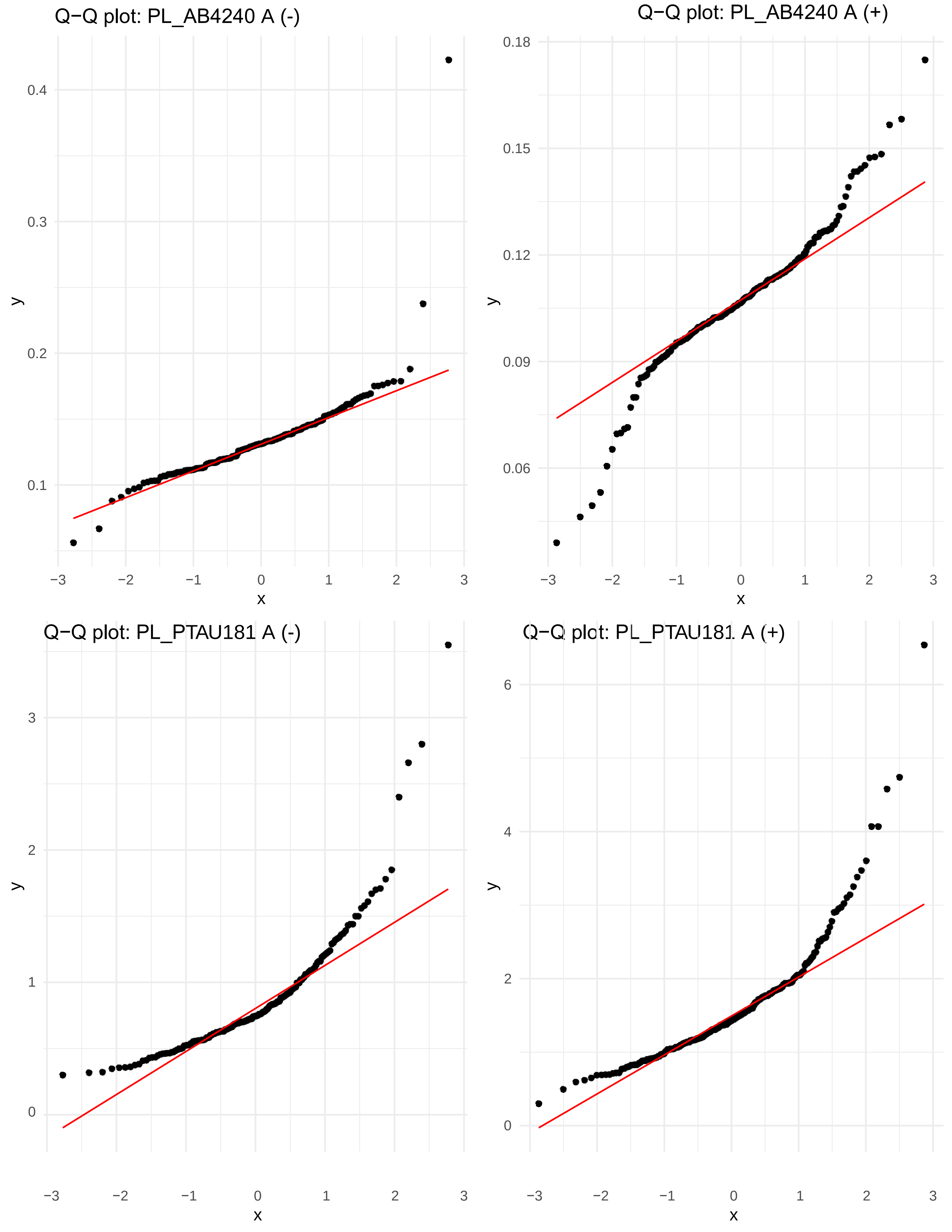
Q−Q plot: PL_AB4240 A (-)
Q−Q plot: PL_AB4240 A (+)
Q−Q plot: PL_PTAU181 A (-)
Q−Q plot: PL_PTAU181 A (+)
x
y

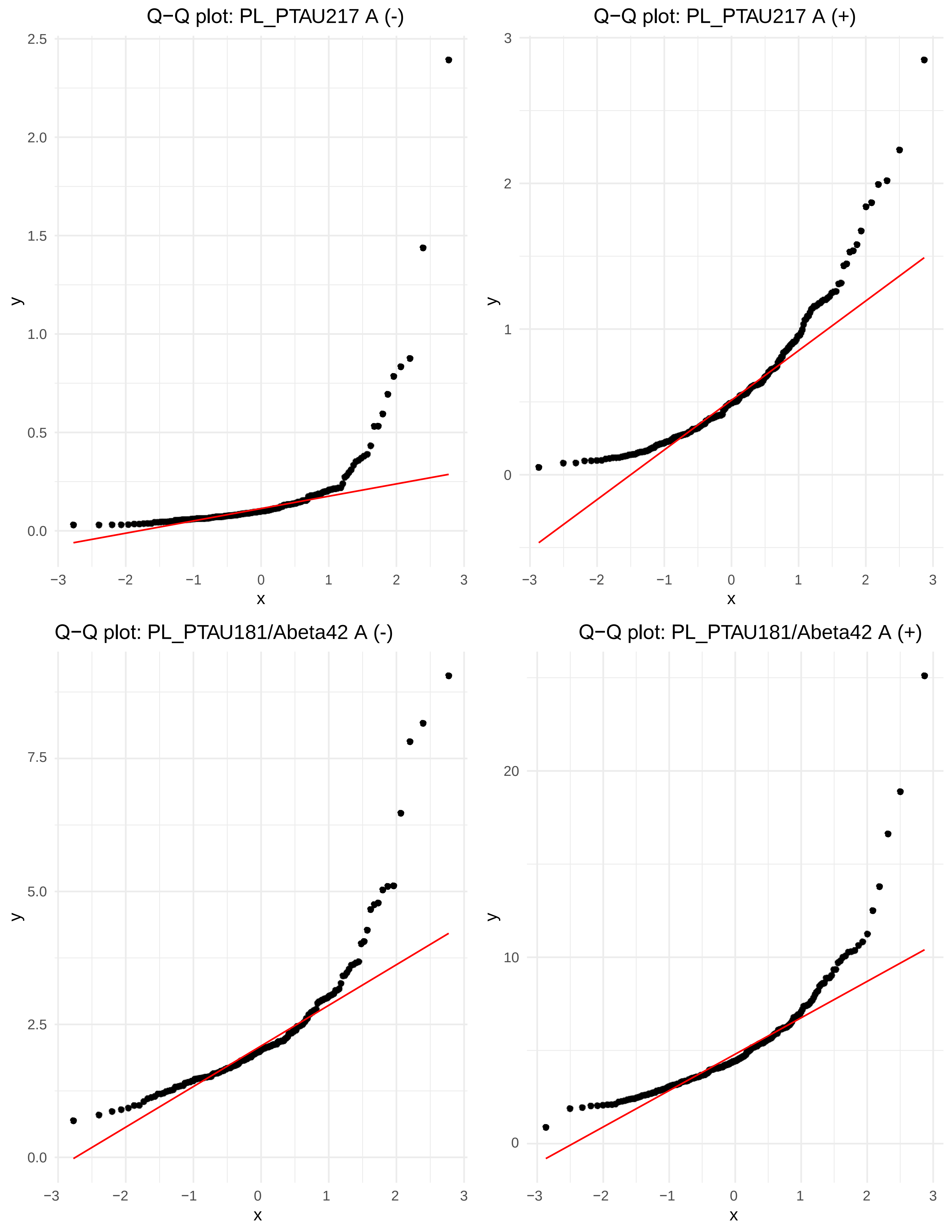
Q−Q plot: PL_PTAU217 A (-)
Q−Q plot: PL_PTAU217 A (+)
Q−Q plot: PL_PTAU181/Abeta42 A (-)
Q−Q plot: PL_PTAU181/Abeta42 A (+)
y
x
0.0
0.5
1.0
1.5
2.0
2.5
0
1
2
3
2.5
5.0
7.5
10
20
−3
−2
−1
0
1
2
3

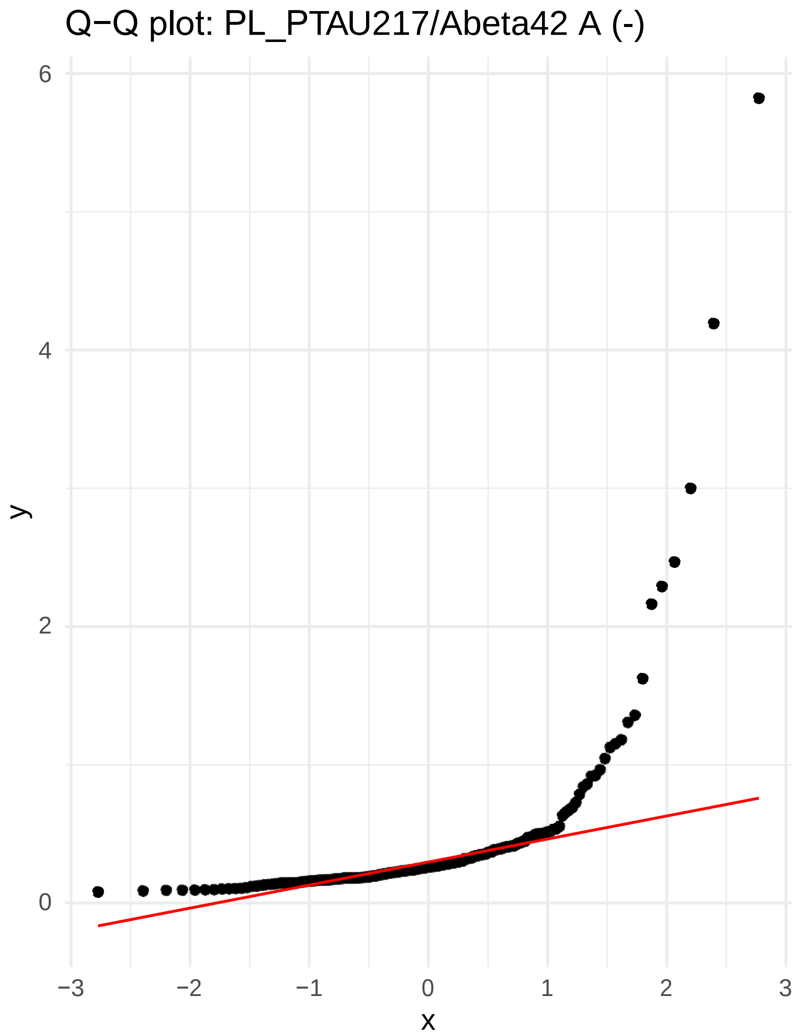
Q−Q plot: PL_PTAU217/Abeta42 A (-)
y
x
0
2
4
6
−3
−2
−1
0
1
2
3

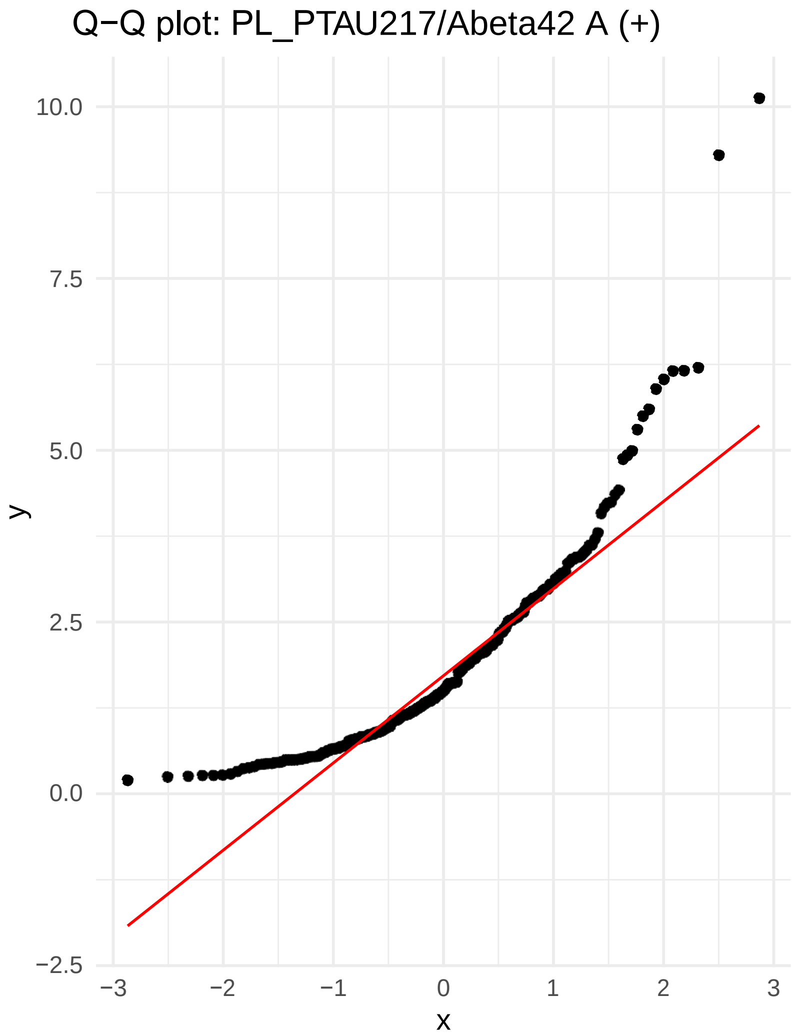
Q−Q plot: PL_PTAU217/Abeta42 A (+)
y
x
−2.5
0.0
2.5
5.0
7.5
10.0
−3
−2
−1
0
1
2
3

**Q-Q plot of the ADNI variables in the Amyloid negative (A(-) and positive A(+) groups**

Legend:
X-axis: Theoretical Quantiles,
Y-axis: Sample Quantiles

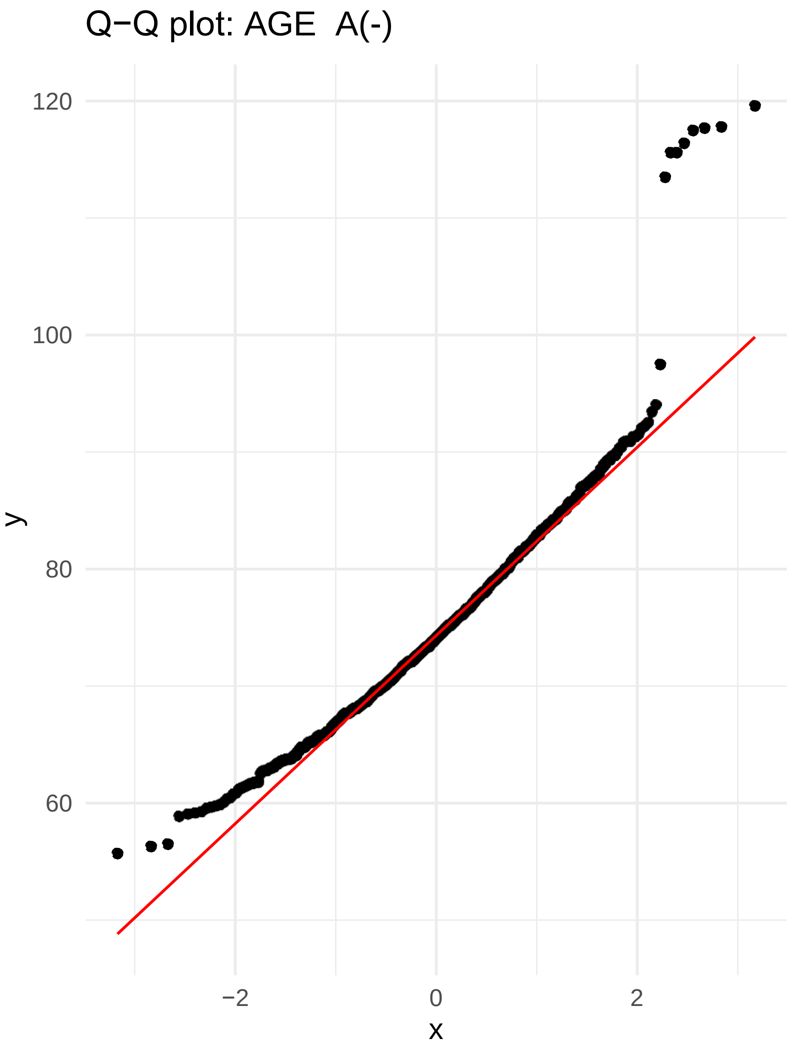


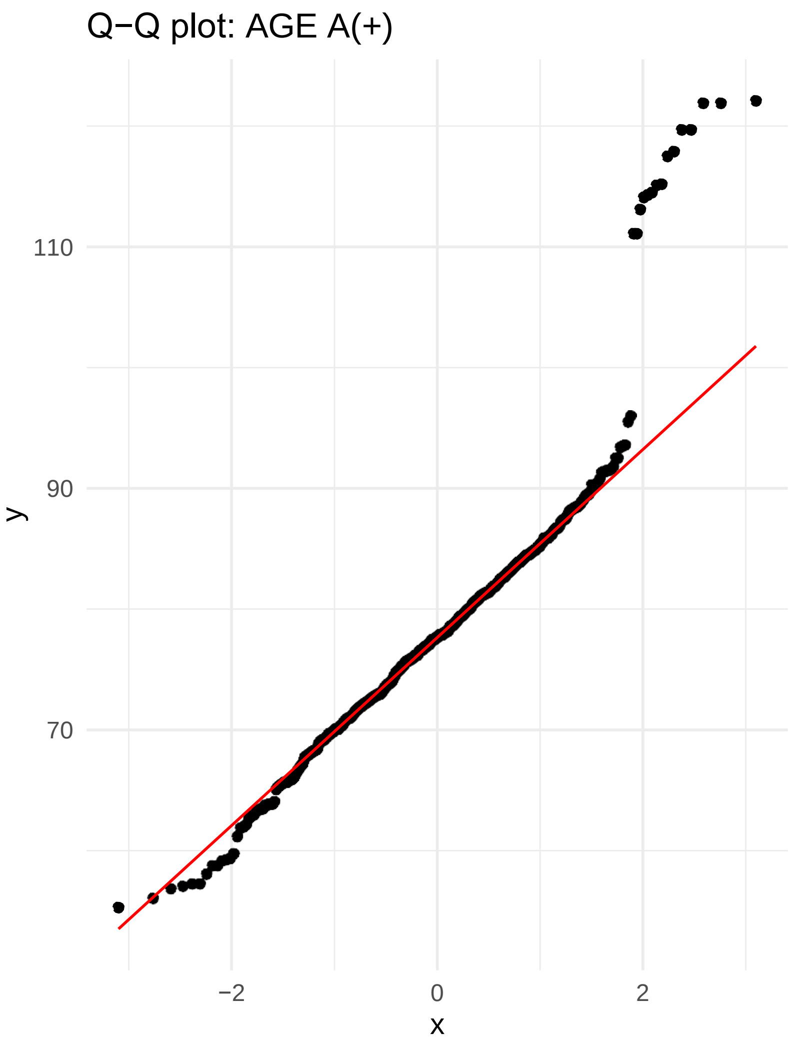


−

−2 0 2
x

−2 0 2
x

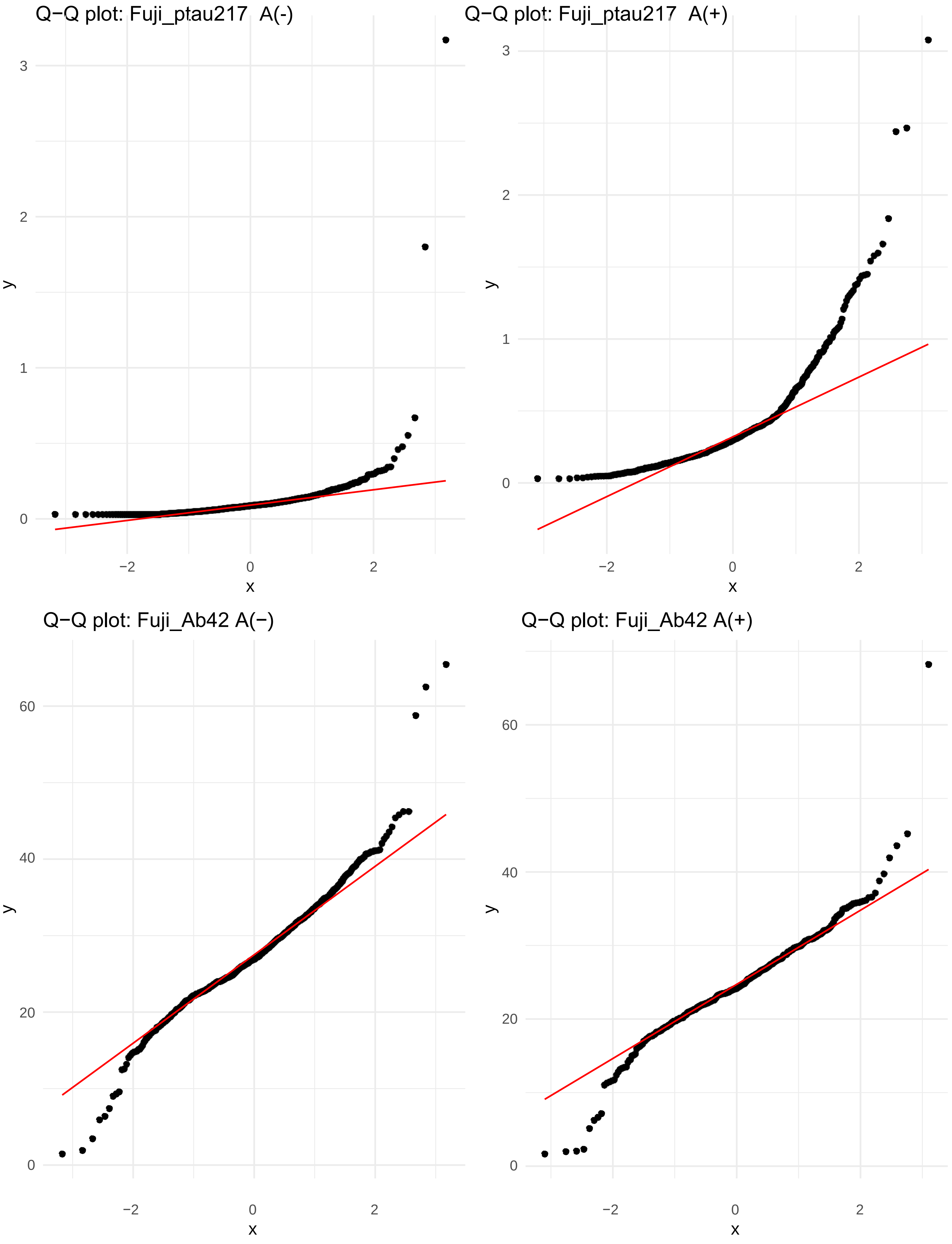
Q−Q plot: Fuji_ptau217 A(-)
Q−Q plot: Fuji_ptau217 A(+)
Q−Q plot: Fuji_Ab42 A(−)
Q−Q plot: Fuji_Ab42 A(+)
y
x
0
1
2
3
−2
0
2
20
40
60

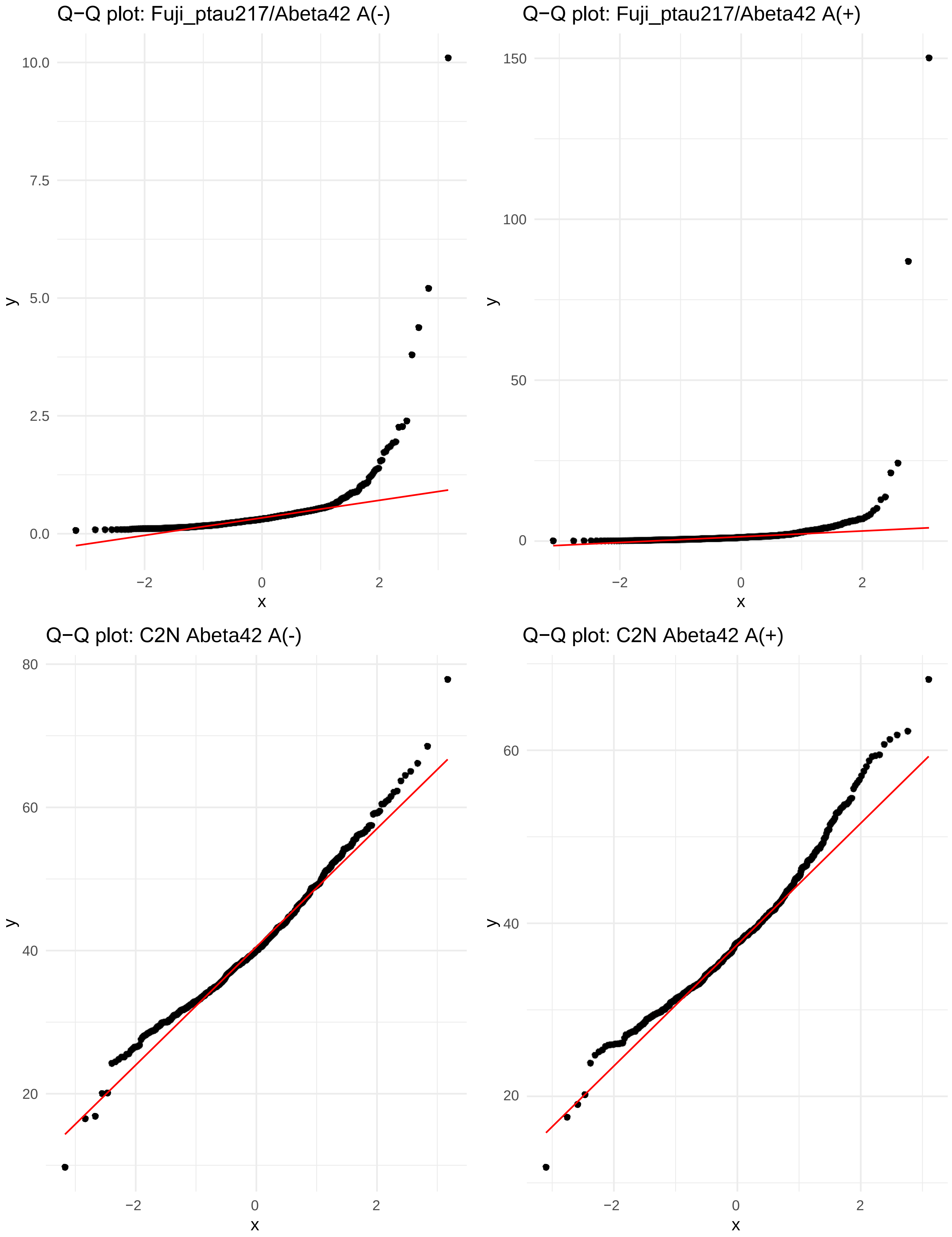
Q−Q plot: Fuji_ptau217/Abeta42 A(-)
Q−Q plot: Fuji_ptau217/Abeta42 A(+)
Q−Q plot: C2N Abeta42 A(-)
Q−Q plot: C2N Abeta42 A(+)
y
x
10.0
7.5
5.0
2.5
0.0
150
100
50
0
80
60
40
20
−2
0
2

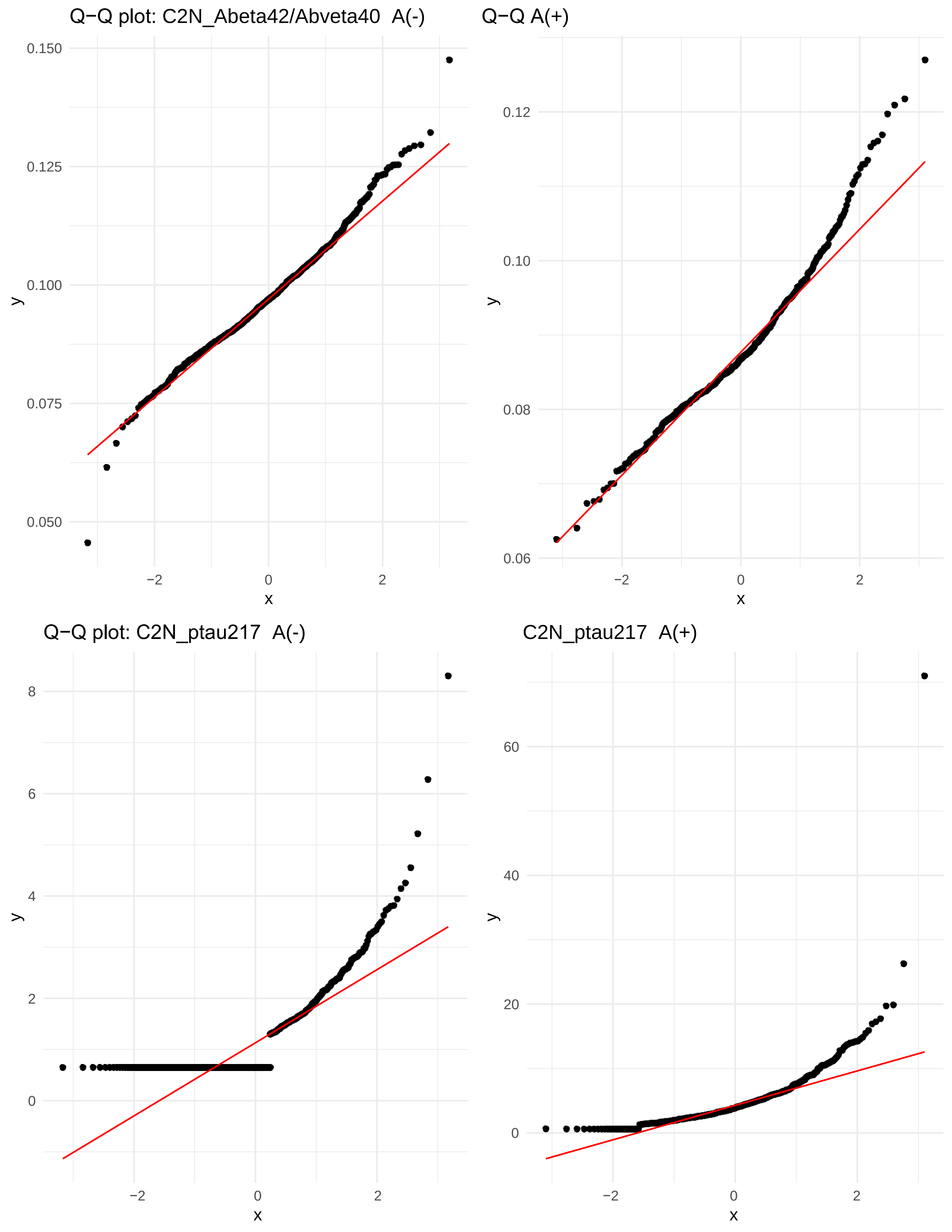
Q−Q plot: C2N_Abeta42/Abveta40  A(-)
Q−Q A(+)
Q−Q plot: C2N_ptau217  A(-)
C2N_ptau217  A(+)
0.150
0.125
0.100
0.075
0.050
0.12
0.10
0.08
0.06
8
6
4
2
0
60
40
20
−2
2
y
x

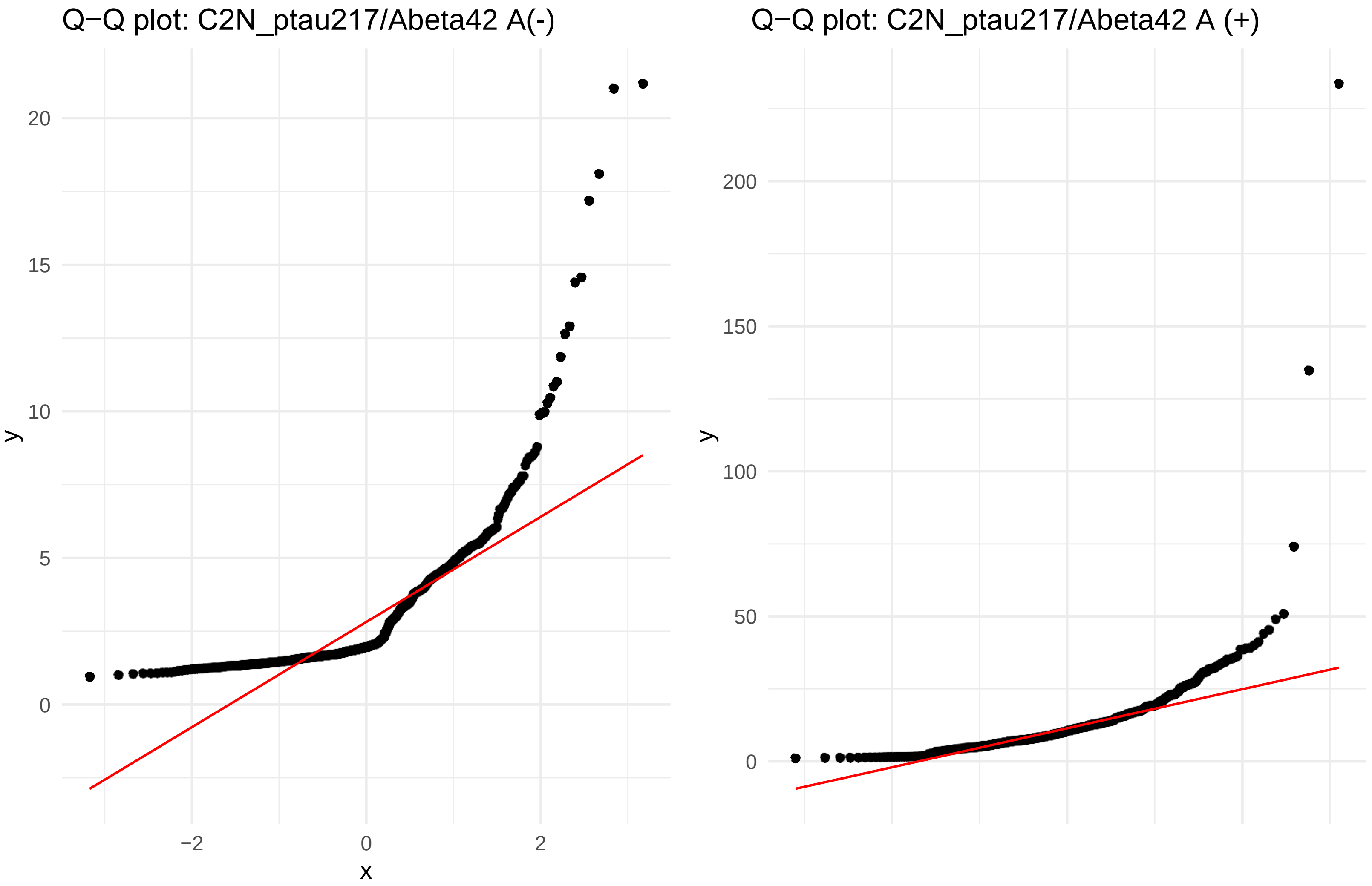
Q−Q plot: C2N_ptau217/Abeta42 A(-)
20
15
10
5
0
y
−2
0
2
x
Q−Q plot: C2N_ptau217/Abeta42 A (+)
200
150
100
50
0
y
